# Lockdown Measures and their Impact on Single- and Two-age-structured Epidemic Model for the COVID-19 Outbreak in Mexico

**DOI:** 10.1101/2020.08.11.20172833

**Authors:** J. Cuevas-Maraver, P. G. Kevrekidis, Q. Y. Chen, G. A. Kevrekidis, Víctor Villalobos-Daniel, Z. Rapti, Y. Drossinos

## Abstract

The role of lockdown measures in mitigating COVID-19 in Mexico is investigated using a comprehensive nonlinear ODE model. The model includes both asymptomatic and presymptomatic populations with the latter leading to sickness (with recovery, hospitalization and death possibilities). We consider situations involving the application of social-distancing and other intervention measures in the time series of interest. We find optimal parametric fits to the time series of deaths (only), as well as to the time series of deaths and cumulative infections. We discuss the merits and disadvantages of each approach, we interpret the parameters of the model and assess the realistic nature of the parameters resulting from the optimization procedure. Importantly, we explore a model involving two sub-populations (younger and older than a specific age), to more accurately reflect the observed impact as concerns symptoms and behavior in different age groups. For definiteness and to separate people that are (typically) in the active workforce, our partition of population is with respect to members younger vs. older than the age of 65. The basic reproduction number of the model is computed for both the single- and the two-population variant. Finally, we consider what would be the impact of partial lockdown (involving only the older population) and full lockdown (involving the entire population) on the number of deaths and cumulative infections.

## I. INTRODUCTION

COVID-19, the disease caused by the novel coronavirus SARS-CoV-2 has, as of August 2020, affected 216 countries and changed the daily lives of billions of people [1]. It has at the same time been the focus of numerous studies, both clinical and mathematical. The study of compartmental models that address the spreading of such epidemics has a time-honored history since the seminal contribution of [2], which by now has been summarized in various reviews [3] and books [4–6]. In recent years, variations to such models focusing on the particularities of coronaviruses have been incorporated, such as the role of asymptomatic carriers of the virus, both as regards earlier CoV examples, such as MERS (for a related example see [7]), and lately in the case of COVID-19 (for a related example see [8, 9]).

Following on some of these more recent developments, the present study focuses on a comprehensive compartmental epidemiological model that takes into account some of the intricacies of COVID-19, while considering its applicability to an urgent and important case example, the country of Mexico. More specifically, the model is an extension of the standard SEIR (Susceptible, Exposed, Infectious, Recovered) model that includes a presymptomatic stage, during which a person experiences no symptoms, but is nevertheless infectious [10]. The proposed mathematical setup also accounts for both asymptomatic infectious and symptomatic infectious individuals. Asymptomatic infectious cases have been found in numerous studies, and reports argue that they may be significantly under-reported [8, 11], which may complicate mitigation efforts such as contact tracing and self-isolation. Those with severe disease symptoms may require lengthy hospitalization, which has strained the health system of many countries [12]. In light of that, the model also includes a compartment describing the hospitalizations. Another distinctive feature of the disease is the heterogeneity with which it manifests in different age groups, especially as it pertains to symptom severity and mortality risk [13, 14]. Other factors, such as preexisting conditions and inter-generational contacts may also play a role [15]. While population age-structure may often be averaged out and de-emphasized in numerous modeling attempts [16], in our model we choose to consider both a single age-group and a two age-group version of the model. The rationale behind this choice is the multifold inhomogeneity in the population of various countries (including our example of interest). Firstly, as mentioned above, the severity in younger people (especially children [17]) is smaller than that in adults. Secondly, older and more vulnerable people may shed more viral particles, thus being more infectious [18]. Thirdly, contacts per day [19] and the contact network itself of older people are different from those of younger people. The partition especially between professionally active (i.e., non-retired) individuals and retirees is important in connection to the above two points, both as regards the differential in average number of contacts of these two groups, and as regards the potential vulnerabilities thereof.

As a case study, we focus on the COVID-19 outbreak in Mexico. While studies for Mexico based on mathematical models exist [20], they differ from ours in several significant ways. Some ignore social-distancing and other mitigation measures [21], others focus on the estimation of the basic reproduction number *R*_0_ and infections using a Bayesian hierarchical model [22], and yet others have since become outdated [23]. Mexico faces a unique challenge, due to the prevalence of COVID-19 risk factors, such as obesity, diabetes and hypertension, among its population [24]. This is reflected in the reported data and our predictive results, which show almost as many fatalities in the < 65 years old group as in the > 65 years old group, even though their populations differ greatly. We feel that these particular features of the Mexican population in conjunction with the large number of infections and especially of deaths in the country warrant an examination through the prism of different age-structured models (e.g., single-age vs. two-age models; in future studies, possibly further partitioning may be of interest) and an assessment of the potential impact of lockdown measures.

Following the formulation of the single-population model and the results obtained through it in section II, we continue with the two age-group model in section III. In each case, we obtain the optimal model parameters in matching the available data regarding deaths, which are considered to be the single most reliable piece of available information. We do discuss the advantages and disadvantages of potentially matching the number of cumulative infections (and the number of deaths). Once the optimal fitting parameters are obtained we assess the impact on both deaths (but also cumulative infections) of immediate lockdown measures in either the case of the entire population or in that of just the older age group. The prediction of the model is that thousands of deaths may be avoided in just a single month alone, should such measures be imposed effective immediately. In section IV we summarize our conclusions and present ideas for future investigation.

## II. SINGLE-POPULATION MODEL

### A. Equations

In the model presented herein, we modify somewhat the setup of the earlier work of a subset of the present authors [25], by incorporating the effect of presymptomatic individuals. More concretely, we start with a susceptible (S) population that can become exposed (E) to the SARS-CoV-2 virus upon interaction with three categories of already infected and infectious individuals: (a) the presymptomatic (P), individuals who are infected, infectious, and eventually will develop symptoms; (b) the asymptomatic (A), individuals who are infected, infectious, and will not develop (clinical) symptoms; and (c) the symptomatically infected/sick (I) population members carrying the virus (infectious). Upon such interaction, the susceptible become exposed to the virus.

Once a member of the population becomes exposed (E), a latent period (*τ*_*l*_ = 1*/σ*_1_) of the virus follows (expected to be in the vicinity of 3 days [26]), during which the exposed population is infected but not infectious. After this period, we assume that the host can naturally be partitioned to either asymptomatic (A) or presymptomatic (P). The fraction of the former is *ϕ*, while of the latter 1 − *ϕ*. While both A and P play a role (along with the infected I) in further transmitting the virus, and indeed A have been argued to play a crucial role [10, 11], it is only P that will present symptoms after an additional time scale, the preclinical period *τ*_*p*_ = 1*/σ*_2_. The incubation period, i.e., the period from infection to the development of (clinical) symptoms, for this partition *τ*_inc_ = *τ*_*l*_ + *τ*_*p*_ = 1*/σ*_1_ + 1*/σ*_2_ is of the order of 5 days [26], and represents the time till the onset of symptoms. It is important to highlight here the relevance of including the presymptomatic population P. Individuals that go down the symptomatic infection path, once they develop symptoms, they will (typically) self-isolate and will interact with fewer other individuals. However, the symptomatic path does not immediately start with symptoms [8]. There is a period when the individual is infectious but does not present symptoms, although they will develop after this preclinical period. During this preclinical time interval, individuals still maintain their regular activity and hence are significantly more impactful towards producing new infections given their “normal” number of contacts.

From there on, the asymptomatics A continue as if nothing happened, given that they have minimal or no symptoms. Their path is only towards recovery with a characteristic rate *M*_*AR*_, or with a time scale representing the asymptomatic infectious period 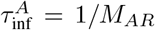, typically expected to be on the order of 7 days. We distinguish those recovered from asymptomatics (who cannot be directly monitored, unless extensive testing is performed in a community) from those coming from a path of symptoms/sickness (who can be –at least partially– monitored), by denoting those recovered from A as A_*R*_.

The path of the presymptomatics P, after becoming infected, is more complicated. Indeed, these may still recover (compartment *R*) without the need for hospitalization (compartment *H*) and without severe manifestation of symptoms (going through class *I*). However, in a fraction *γ* of the cases hospitalization is needed. Recovery or hospitalization in the model is associated with a time scale 1*/M*, the symptomatic infectious period 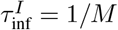. Subsequent steps involve a fraction *ω* of the hospitalized that die, over a time scale 1*/ψ* and a fraction 1 − *ω* that recover over a time scale 1*/χ*. The above offers, in principle, a complete description of the modeled quantities within our system. We should note that the transmission rates *β* refer to the coefficients of interaction between A (or P) and S, as well as I and S, leading to new infections; these are, respectively, *β*_*SA*_ and *β*_*SI*_. We note here, that we somewhat abuse notation as far as *β*’s are concerned: we consider the transmission rate 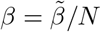, where *N* is the total population. What we report in the tables that follow is actually 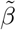. In terms of the relevant time scales, the infectious period associated with symptomatically infected is 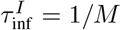, with asymptomatic individuals 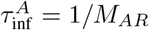, whereas the infectious period associated with presymptomatic individuals is 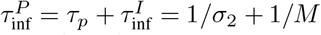.

The transcription of the above pathways in equations leads to the following ODEs:

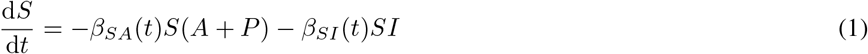

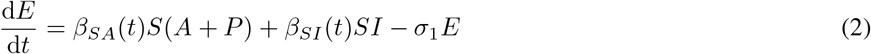

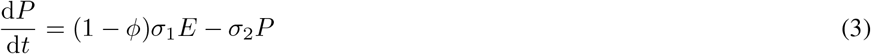

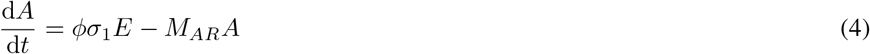

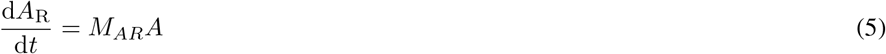

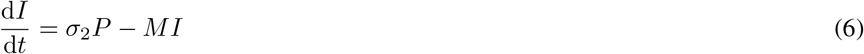

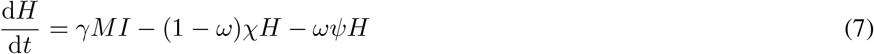

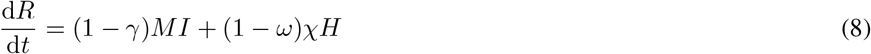

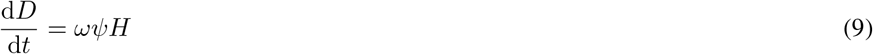

A schematic diagram of the model is shown in Fig. 1.

**FIG. 1.**
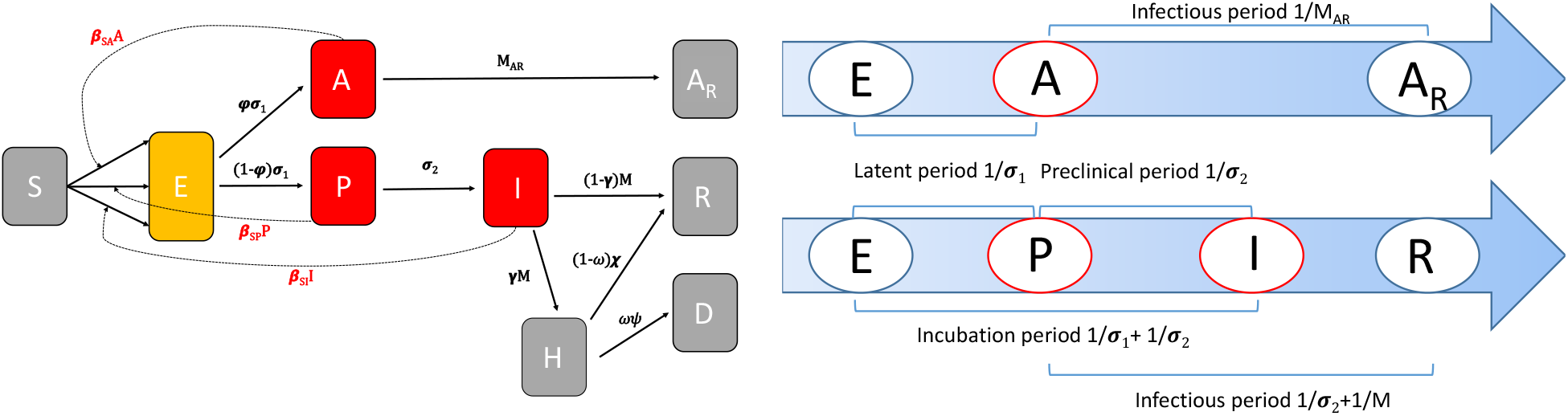
Schematic diagram of the single-population model (left panel) and a diagram of the two disease-progression pathways: the asymptomatic and the symptomatic (right panel, based on a variation of a diagram in [8], adapted to the compartments and time scales of our model).

We will consider the data for Mexico, with a total population of 127,575,528 people (in 2019) and more than 430,000 confirmed cases and 47,000 deaths by the beginning of August 2020. Data were taken from “Dirección General de Epidemiología (DGE)” of “Gobierno de México” [27]. These data have the particularity of including every clinical case, from which we retrieve three basic pieces of information: the date symptoms start, the death date (if applicable), and the patient’s age. The data were updated on a daily basis. As the way of measuring always implies an underestimation of the number of cases and deaths in the days close to the report’s date (especially because of the delay in death communications to the DGE), we performed fits up to dates about 20 days from the report date (i.e., the report date is July 29 and fits are performed until July 10).

Our principal diagnostic quantities to obtain the optimal model parameters will be the time series for deaths *D*(*t*). In addition, we will also monitor the cumulative infections, defined in the realm of the present model as *C*(*t*) = *I*(*t*) + *H*(*t*) + *R*(*t*) + *D*(*t*), i.e., the sum of the individuals going through the network pathway involving the symptomatically infected. We take *t* = 0 as March 22, and fit data up to July 10. This reflects our effort to be (in terms of the total numbers of both diagnostics) well within the “well mixed” regime from the beginning of the infection where the ODEs of interest are expected to be relevant.

We are particularly interested in the effect of non-pharmaceutical intervention strategies on the spreading and development of the disease. It is, thus, important to specify the actual intervention measures and their timing, along with proposed subsequent hypothetical intervention scenarios and their effect on the *predicted* development of the epidemic. In the case of interest, the country of Mexico, on March 22 a partial lockdown was imposed [28] in Mexico city, whereby, e.g., bars, nightclubs, movie theaters, and museums were closed. As mentioned, this is the initial day of our fitting. On April 21 (*t*_*q*_ = 30), the country entered Phase 3 of its contingency plan, whereby instead of the virus leading to community transmission (as in Phase 2), the entire country was in a state of epidemic: this implied stricter health protocols. The associated intervention measures primarily restricted human mobility by requiring social distancing: they will be referred as such in the following. Subsequent to these actual measures, we shall consider a number of scenarios of more severe measures, e.g., complete lockdown and the requirement that face masks be worn, which may have been imposed on August 10 (*t*_*L*_ = 141).

Intervention strategies render the transmission rates time dependent. Their effect on the overall transmission rate *β* may be estimated by considering biological and physical properties of expelled respiratory droplets, the carriers of the pathogens, and specifically of SARS-CoV-2. The transmission rate is usually written as *β* = *cp*, with *c* the average number of contacts per day a susceptible has with an infectious individual and *p* the disease-transmission probability. For pathogen transmission via infectious respiratory droplets, be they airborne (airborne transmission) or settled (contact transmission), the airborne transmission rate has been expressed as, cf. Refs. [29] and [30]

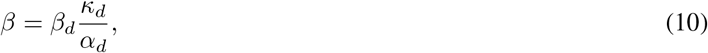

where *κ*_*d*_ is the respiratory droplet emission rate (viral shedding) by e.g. breathing, speaking, coughing, sneezing, singing, *α*_*d*_ is the airborne, infectious droplet effective removal rate, by e.g. gravitational settling, ambient airflow, pathogen inactivation, and *β*_*d*_ the transmission rate per deposited respiratory droplet. As we will use Eq. (10) to *estimate* how the overall transmission rate changes we will neglect its complex dependence on droplet diameter. The droplet transmission rate *β*_*d*_ depends on the number of effective contacts a susceptible has with other individuals, the number of pathogens contained in an infectious droplet (its pathogen load), and the virus transmission probability per inhaled/deposited droplet.

Social distancing or other lockdown measures that restrict human mobility decrease the average number of daily contacts, and possibly the average duration of contact, thereby decreasing *β*_*d*_ (and consequently *β*). We denote this fractional decrease, which arises from the social-distancing measures imposed on April 21, as *η*_*SA*_ and *η*_*SI*_. Another common intervention measure is the use of personal protective equipment, e.g., surgical face masks. Milton et al. (2013) [31] argued that their use produced a 3.4-fold reduction in viral aerosol shedding. Face masks remove expelled respiratory droplets by filtering them and they modify the expelled air flow, with consequential effects on droplet transport and dispersion in the environment and their airborne lifetime (thus, their removal rate). The oversll effect of wearing surgical face masks leads primarily to a decrease of viral shedding (decreased droplet generation rate *κ*_*d*_). Their effect is difficult to estimate: we follow [32] to estimate that their use (if the whole population used them continuously and correctly fitted) would decrease the overall transmission rate to *≈* 0.5*β*.

Further intervention measures in the scenarios that we consider are modeled by the parameter *ζ*, Eq. (11): we set the asymptomatic transmission rate, as discussed following Eq. (11), to be *η*_*SA*_*ζβ*_*SA*_. This parameter incorporates the effect of additional (to those imposed on April 21) intervention measures, including the requirement that face mask be worn. We varied *ζ* from 1.0 to 0.5. Thus, whereas *η* simulates actual intervention measures, *ζ* simulates potential intervention scenarios.

These features should be kept in mind, as we aim not only to capture the current trend of the pandemic, but also to suggest mitigation strategies that may reduce cumulative infections, as well as COVID-19 induced fatalities in the time series of Mexico. We explore the impact of *additional* intervention measures to mitigate the spread of the infection by the following time dependence of the transmission rates, *β*:

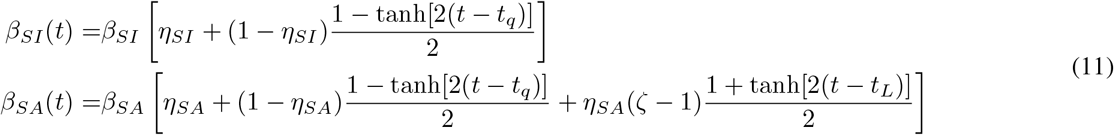

with 0 < *ζ* ≤ 1, as argued, and *t*_*q*_, *t*_*L*_ previously specified. For *ζ* = 1, Eqs. (11) reduce to the equations modelling the decrease in the number of personal contacts when a light form of social distancing was imposed on August 21 (no lockdown, no requirement to wear face masks). Figure 2 shows their time-dependence. The first jump of *β*_*SI*_ and *β*_*SA*_, the transmission rates associated with infected and presymptomatic-asymptomatic virus carriers, occurs at *t*_*q*_. The next jump occurs at *t*_*L*_: it is assumed to occur only for the presymptomatic or asymptomatic carriers of the virus through the additional hypothetical restrictions at *t* = *t*_*L*_. The transmission rate associated with infected individuals, *β*_*SI*_, exhibits only one decrease as we consider that symptomatic individuals are completely isolated (self isolation).

**FIG. 2.**
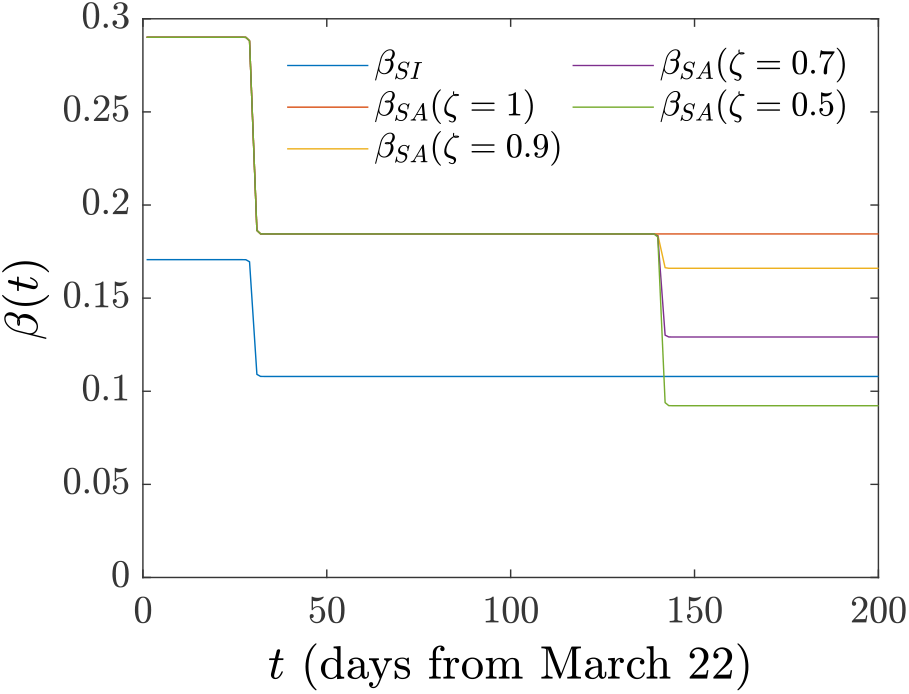
Single-population model. Time dependence of the transmission ratres *β*_*SI*_ and *β*_*SA*_. For the asymptomatic carriers of the virus the transmission rate initially decreases due to social distancing as reflected in *η*_*SA*_. It further decreases by a factor *ζ* (each corresponding to a different intervention strategy) reflecting assumed additional intervention measures, like obligatory wearing of face masks or additional mobility restrictions (lockdown). For the infected population instead, we assume that *β*_*SI*_ decreases only once due to social distancing actually imposed on April 21, (*η*_*SI*_): it does not decrease further, given the self-isolation of infected individuals with symptoms.

## B. Results

We obtained the optimal model parameters by minimizing the distance between model-predicted time series and the corresponding time series obtained from the Government of Mexico [27]. The distance was specified by the following combination of Euclidean (*l*^2^) norms

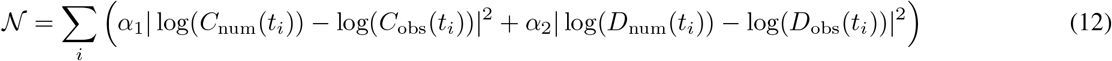

where the index “i” identifies the point in the time series, the subscript “num” refers to the predicted time series, the subscript “obs” to the observed time series. We performed 2,000 optimizations with an initial guess for each parameter uniformly sampled within a pre-specified range (see Table I for the variation range). The upper and lower limits of the variation ranges were used as boundaries in the constrained minimization algorithm (implemented in Matlab via `fmincon` function). The parameter ranges were determined from epidemiological information. Moreover, we considered *σ*_1_ and *σ*_2_ (via the appropriate variation ranges) such that 5 ≤ 1*/σ*_1_ + 1*/σ*_2_ ≤ 6, a constraint that arises from the SARS-CoV-2 [26] observation that its incubation period *τ*_inc_ = 1*/σ*_1_ + 1*/σ*_2_ is roughly 5 ≤ *τ*_inc_≤ 6 days from the start of exposure. We note that at the initial time of the model fitting, the number of exposed, asymptomatic, and presymptomatic is not known. We thus optimized their ratio to the initially infected *I*(0), a number that was obtained by subtracting the officially reported number of deaths, recovered, and hospitalized from the (reported) number of cases. Lastly, due to parameter identifiability, discussed below, we fixed *ω* = 0.1292. This was the value of *ω* identified in our optimization computations when *ω* was kept as a free parameter. To corroborate whether this was a “reasonable estimate” of the fraction of fatalities we examined the fraction of fatalities over hospitalizations incurred within the data for the total population. We found the latter number to be *ω* = 0.1234, i.e., remarkably close to the above estimate, hence we opted to fix *ω* to the above value of 0.1292. The sensitivity of the predicted model parameters (with the previously mentioned constraints) when the ratio of *β*_*SA*_ to *β*_*SI*_ is allowed to vary within a specified interval ([0.2, 2]) and parameter-identifiability issues are discussed in Section II C.

We considered two cases: *α*_1_ = 0, *α*_2_ = 1, results displayed in Fig. 3 with corresponding optimal model parameters shown in Table I, and *α*_1_ = *α*_2_ = 0.5, results shown in Fig. 4. Table I presents both the median and the interquantile range for each parameter (as they arise from the 2,000 optimizations), as well as the range of variation of the initial guesses of the model parameter and initial conditions. The last three entries give the ratio of exposed, asymptomatic, and presymptomatic individuals to the infected at the initial time of our fitting. The rationalization for the two choices of the norm is as follows. In the former case (*α*_1_ = 0, *α*_2_ = 1), we take the view that the only “ground truth” data are those of the deaths; in fact, even these can be under-estimated (deaths with “suspicion” of COVID-19, but no definitive test) or –perhaps less likely– over-estimated (deaths attributed to COVID-19 without explicit testing). Indeed, as is highlighted in [34], “the actual total death toll from COVID-19 is likely to be higher than the number of confirmed deaths – this is due to limited testing and problems in the attribution of the cause of death”. Nevertheless, here we assume that this is the most well-defined piece of data, as is generally expected to be the case. On the other hand, infections are more broadly expected to be under-reported. This is because many of the cases with symptoms do not get to be serious enough to lead to hospitalization or to be reported. In that light, it is expected that assuming the deaths as ground truth, we should expect to find a significant over-estimation of the number of infections (we return to this point below). If, on the other hand, we “force” the model to match the current reporting of infections as well, we will end up with a better approximation “on average” to both curves but with a potential adverse by-product in the resulting number of deaths. By “force” we mean that our minimization scheme tries to minimize the distance of *both* observed deaths and observed cumulative infections, instead of (as above) considering only the deaths as ground truth. Issues of measurement errors in the setting of cumulative incidences have been discussed, in e.g., in [33], further supporting the choice *α*_1_ = 0, *α*_2_ = 1. We will discuss this further in the context of our results below.

**FIG. 3.**
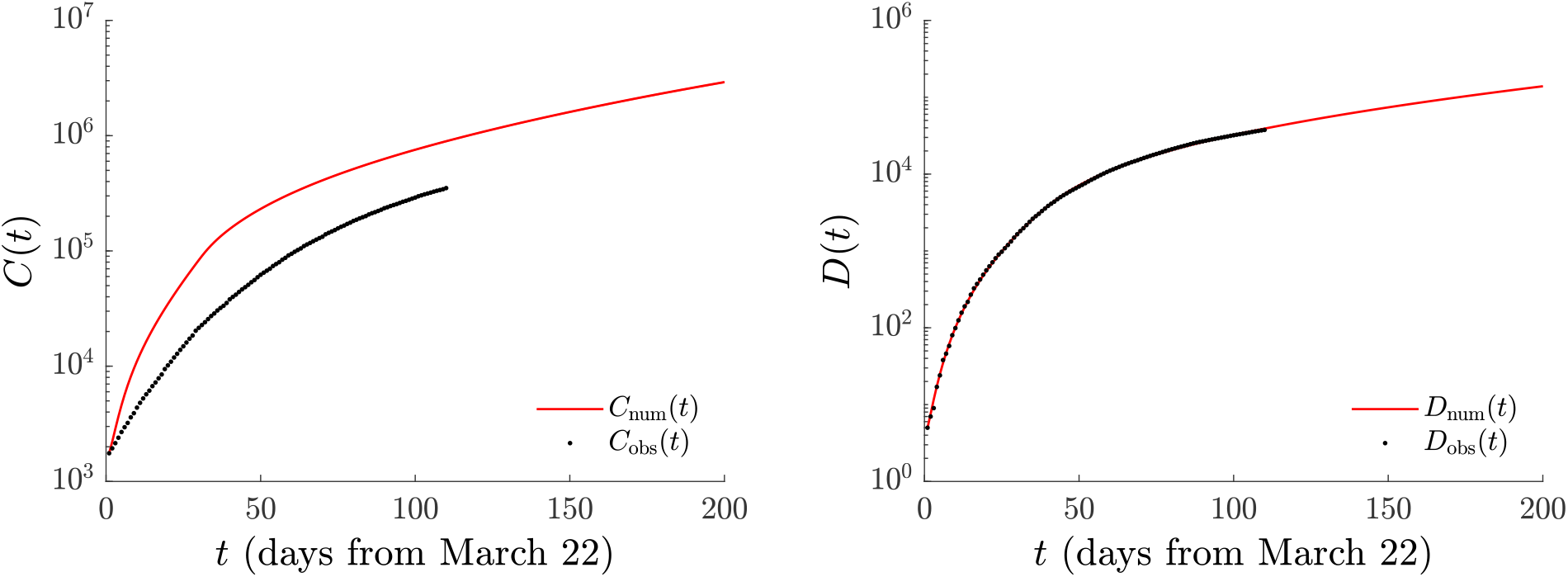
Single-population model. Number of cases *C*(*t*) (left) and deaths *D*(*t*) (right) found by minimizing the Euclidean norm (12) with *α*_1_ = 0, *α*_2_ = 1. The fit used data until July 10. The (optimized) model predictions are shown by the solid (red) line, while the official time series [27] is given by (black) dots. The shaded area, corresponding to 2000 simulations with parameters chosen within the interquartile ranges, is too narrow to be visible in the scale of the plot.

**TABLE I.** Single-population model. Optimal parameters (median and interquantile range). Euclidean-norm minimization with *α*_1_ = 0, *α*_2_ = 1. Fixed fraction of hospitalized individuals who died *ω* = 0.1292 (see text for justification). Variation range used in the optimization algorithm (initial parameter and initial-condition guesses were uniformly sampled within the ranges shown).

| Parameter | Symbol | Median (interquartile range) | Initial value |
| --- | --- | --- | --- |
| Population | $N$ | 127,575,528 | |
| Initial populations | $(I_0; H_0; R_0; A_{R_0}; D_0)^a$ | (1,577; 171; 4; 0; 5) | |
| Transmission rate [per day] | $\beta_{SI}^b$ | 0.1707 (0.1592–0.1844) | $c \in U[0, 1]$ |
| Transmission rate [per day] | $\beta_{SA}^b$ | 0.2901 (0.2773–0.3021) | $c \in U[0, 1]$ |
| Social distancing effect | $\eta_{SI}$ | 0.6325 (0.6004–0.6623) | $c \in U[0, 1]$ |
| Social distancing effect | $\eta_{SA}$ | 0.6360 (0.6074–0.6648) | $c \in U[0, 1]$ |
| Latent period [days] $\tau_l$ | $1/\sigma_1$ | 2.9692 (2.9661–2.9716) | $1/k, k \in U[2.75, 3.25]$ |
| Preclinical period [days] $\tau_p$ | $1/\sigma_2$ | 2.4545 (2.4524–2.4563) | $1/k, k \in U[2.25, 2.75]$ |
| A/P partitioning | $\phi$ | 0.1922 (0.1796–0.2038) | $c \in U[0, 1]$ |
| Infectivity period (I) [days] $\tau_{inf}^I$ | $1/M$ | 5.5780 (5.4718–5.6639) | $1/k, k \in U[3, 14]$ |
| Infectivity period (A) [days] $\tau_{inf}^A$ | $1/M_{AR}$ | 6.8822 (6.8210–6.9363) | $1/k, k \in U[5, 14]$ |
| Conversion fraction (I to H, R) | $\gamma$ | 0.5409 (0.5349–0.5479) | $c \in U[0.25, 0.75]$ |
| Recovery period(H to R) [days] | $1/\chi$ | 9.3753 (9.2915–9.4906) | $1/k, k \in U[5, 15]$ |
| H to D period [days] | $1/\psi$ | 12.0078 (11.9828–12.0404) | $1/k, k \in U[5, 15]$ |
| Initial population fraction (E) | $E(0)/I(0)$ | 2.1422 (2.0626–2.2537) | $c \in U[1, 3]$ |
| Initial population fraction (A) | $A(0)/I(0)$ | 2.0964 (2.0198–2.2061) | $c \in U[1, 3]$ |
| Initial population fraction (P) | $P(0)/I(0)$ | 0.5352 (0.4995–0.5792) | $c \in U[0, 1]$ |

<sup>a</sup> Initial susceptible population $S_0 = N - (I_0 + E_0 + A_0 + P_0 + H_0 + R_0 + A_{R_0} + D_0) \approx N$ .
<sup>b</sup> The reported transmission rates $\beta$ have been multiplied by $N$ . They should be normalized by the initial population when used in ODE model.

**FIG. 4.**
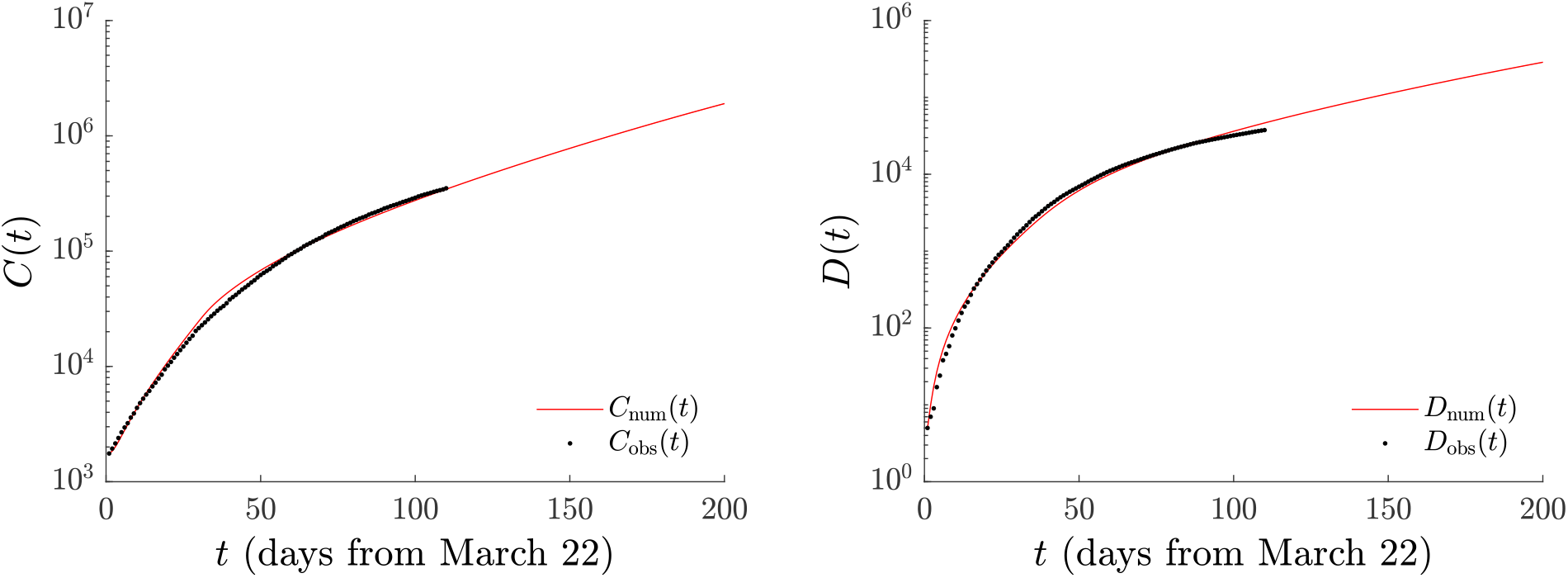
As in Fig. 3, but with *α*_1_ = *α*_2_ = 0.5. The number of cases *C*(*t*) is captured significantly better, but the lower accuracy in capturing *D*(*t*) suggests considerably higher predicted number of deaths in the future. The shaded area, corresponding to 2000 simulations with parameters chosen within the interquartile ranges, is too narrow to be visible in the scale of the plot.

Inspection of Fig. 3 shows that the model provides an excellent fit to the total number of deaths with a considerable over-prediction of the number of infections, presumably (as argued before) because numerous infections are not reported in the official data. It is important to highlight that in this case (*α*_1_ = 0, *α*_2_ = 1) the number of cumulative infections *C*(*t*) is *not* fitted but it is a model output. We note that the significantly higher number of *C*(*t*) is in line with the widely popular, yet hard to quantify, expectation that the true number of infections is significantly higher than officially reported. We will find similar features when we interpret the two-age model predictions.

These results can be compared with Fig. 4 where both deaths and cumulative infections are fitted in the minimization procedure, *α*_1_ = *α*_2_ = 0.5, Here, we note that while the model does a very adequate job in following *C*(*t*), it also does a reasonable job of capturing *D*(*t*). *Nevertheless*, there is a caveat to the latter. A closer observation of the semi-logarithmic scale would lead the astute reader to observe that as the model is trying to juggle the optimization of both time series, it slightly over-predicts deaths early on, slightly under-predicts them at the middle of the time series, and eventually slightly over-predicts again at longer times. This minimization likely predicts a scenario with multiple hundreds of thousands of deaths at the end of the examined evolution that are not warranted by the data trends. For this reason, we will stick to the consideration of the former case of Fig. 3 hereafter. Indeed, we should note that we have tried similar optimization procedures on various similar data sets (stemming from different countries or autonomous regions [25]) and this feature has been generic. Namely, when attempting to fit both the deaths and cumulative infections, the deaths (which we expect to be “less under-reported”) deviate considerably more significantly past the final time of the fit and suggest a lesser predictive ability thereof in the future (than if working with the fatalities dataset alone). It is in that light that we select to only work with the latter set. Doing so has generically led our optimizations to identify a larger number of infections than the ones reported not only for Mexico, but also for other countries and autonomous regions for which our methods have been applied. In that vein, our results are *merely suggestive* of the broadly expressed view of the infections being much higher than the ones reported. It is, thus, not surprising that in [34], it is indicated that “the actual number of cases is likely to be *much* higher than the number of confirmed cases” (with the emphasis being ours).

Indeed, it may appear as somewhat unusual to have data available for *C*(*t*) and choose not to use it. Yet, the careful collection of data seems to increasingly suggest that the number of infections is substantially under-reported in comparison to true infections [35], not only because of asymptomatic carriers of SARS-CoV-2, but also due to individuals that did not seek medical care. Indeed, the relevant study of the Centers for Disease Control and Prevention in the US finds an under-reporting of infections by a factor of 2 to 13 times, which is quite well within the margin of our observations. It appears to us natural to expect it would be *unlikely* for the uncertainty in deaths to be anywhere near such multiplicative factors. In short, we do not feel that selecting to consider *D*(*t*) as the ground truth is an aspect raising doubts about the validity of the model, but rather it is a finding posing the question about the potential under-reporting of the cumulative infections, a general trend also applicable to the case of Mexico. We will notice similar features in the case of the two-age model.

Before we discuss the implications of mitigation strategies, let us briefly comment on the optimal model parameter (median) values, as summarized in Table I. The latent period 1*/σ*_1_ is indeed found to be in the vicinity of 3 days (2.97), while the total incubation period is comfortably within the prescribed interval of 5-6 days (1*/σ*_1_ + 1*/σ*_2_ *≈* 5.42). The time scale of recovery for asymptomatics is a little under 7 days as expected (1*/M*_*AR*_ = 6.88), while the time scale of going from symptomatic infected to hospitalization is close to 6 days which is also fairly reasonable (1*/M* = 5.58). The model predicts the fraction of asymptomatics to be about 20% (*ϕ* = 0.19) which is smaller than reported elsewhere [11]. However, such results should be taken with a grain of salt due to parameter identifiability [36]. In particular, a systematic analysis of the model along the lines of [41] suggests that *ϕ* is not itself an identifiable parameter, but rather the product of *ϕ* with *β*’s might be. We will provide more details on this point in the subsection II C that follows.

Interestingly, the model predicts that roughly 55% of those symptomatically infected need hospitalization, while the rest recover. The average time scale of recovery upon hospitalization is about 9.4 days (1*/χ*), while that of death (1*/ψ*) is found to optimally be near 12 days. However, here too we should highlight that *ω* and *ψ* are not independently identifiable and neither is (1 − *ω*) and *χ*.

Our aim is now to explore the impact of additional, scenario-induced mitigation measures that reduce the obtained optimal transmission rate *β*_*SA*_ by a factor *ζ <* 1, cf. Eq. (11) (*ζ* = 1 gives the optimization result without additional hypothetical measures). Table II summarizes the results of Fig. 5, in which the predictions for *ζ* = 0.9, 0.7, 0.5 are displayed. It can be seen that a decrease of *ηβ* by a factor between 0.9 and 0.5 could have a *catalytic* result as concerns the predictions of the model both for deaths and cumulative infections. In particular, a decrease of *ζ* by a factor of even as little as 0.9 would reduce the number of deaths by more than 1,500, only within the time frame between August 10 and September 10, while a decrease by a factor of 1*/*2 is predicted to save close to 6,100 lives in this interval alone. The corresponding effect on the number of infections is perhaps even more striking (also noting that many of these infections could lead to fatalities at a later stage). In particular, a decrease of *ζ* to 0.9 leads to about 75,000 fewer infections, while a decrease by 1*/*2 would lead to about 288,000 fewer infections, again just within this 31-day period. One can clearly see the significant potential impact of further lockdown measures (and the requirement that face masks be worn), a feature that may be worthwhile to factor into further public health considerations.

**TABLE II.** Single-population model. Predictions for *C* and *D* at September 10 if additional lockdown measures had been applied on August 10.

| | $\zeta = 1$ | $\zeta = 0.9$ | $\zeta = 0.7$ | $\zeta = 0.5$ |
| --- | --- | --- | --- | --- |
| $C$ (total) | 2,119,475 (2,074,088–2,154,804) | 2,044,226 (2,003,756–2,078,877) | 1,921,479 (1,886,812–1,955,272) | 1,829,307 (1,793,600–1,861,605) |
| $C$ (from Aug. 10) | 695,412 (679,573–708,702) | 620,571 (607,195–633,057) | 500,540 (481,878–515,504) | 407,116 (385,326–427,064) |
| $D$ (total) | 99,184 (98,760–99,489) | 97,656 (97,218–97,997) | 95,051 (94,525–95,483) | 92,870 (92,261–93,422) |
| $D$ (from Aug. 10) | 34,327 (34,061–34,513) | 32,814 (32,516–33,034) | 30,195 (29,759–30,572) | 28,030 (27,458–28,545) |

**FIG. 5.**
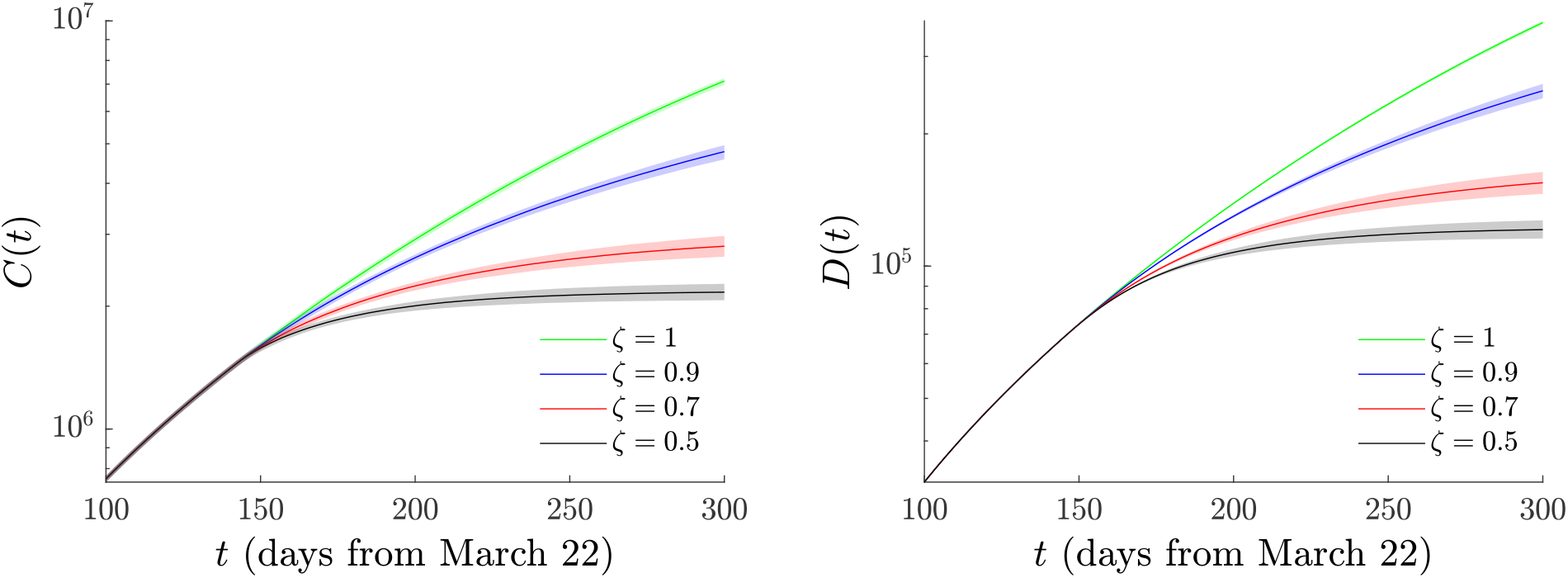
Single-population model. Evolution of the number of cases *C*(*t*) (left) and of deaths *D*(*t*) (right) for different *ζ*’s if additional lockdown measures had been applied on August 10 (*t* = *t*_*L*_ = 141). Notice the significant curbing of the pandemic as a result of such interventions, especially the long-term effects for *ζ* = 0.7 and *ζ* = 0.5.

As our final comment on the single-population variant of the model, we note that an important quantity in epidemiological models of this type is the basic reproduction number *R*_0_; see, e.g., [3]. It is the number of expected new infections (secondary infections) arising from a single infectious individual in a population where all subjects are susceptible. Using the next-generation approach [37], we determine the basic reproduction number as a function of model parameters and the initial susceptible popu-lation (see Appendix A 1 for details) to be

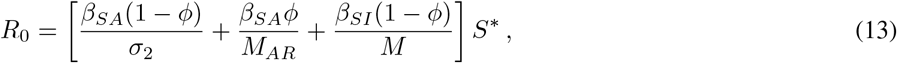

with *S*^*^ the initial susceptible population, *S*^*^ = 1 as parameters *β* are normalized by *N*. For the parameters in Table I, *R*_0_ = 1.73 (median; interquartile range 1.71 − 1.75). We also calculated the effective reproduction number, i.e., the number of cases generated in the current state of population (which does not have to be necessarily the uninfected state), at the beginning of social distancing measures (*t* = *t*_*q*_) using *β*_*SA*_(*t*_*q*_) and *β*_*SI*_ (*t*_*q*_) in Eq. (13). We found *R*_*e*_ = 1.41 (median; interquartile range 1.40 − 1.42) which suggests that the pandemic was not mitigated by the intervention measures. The spreading of the infection is predicted to start to decrease for *R*_*e*_ < 1: this requires that *ζ* be smaller than 0.84 (median; interquartile range 0.82 − 0.86). We obtained this value by setting the right hand side of Eq. (13) to 1, after replacing *β*_*SA*_ → *η*_*SA*_*ζβ*_*SA*_ (as a result of the proposed additional lockdown measures), keeping the infected transmission rate to be *η*_*SI*_*β*_*SI*_. The resulting equation is an algebraic equation for *ζ* whose solution yields the reported *ζ*. The calculated effective reproduction number explains why among the case examples we considered, those with *ζ* = 0.7 and 0.5 present a significant decrease in the number of deaths not only immediately (i.e., within the interval of August 10 to September 10) but also over the longer-scale prediction of Fig. 3.

### C. Sensitivity Analysis to Parameter Variations and Initial Conditions, and Identifiability Issues

A very natural question regarding our optimization routine and its findings concerns its sensitivity towards parametric variations, as well as variations of initial conditions, and the associated confidence intervals. We considered various methods, among the many that exist in the literature, see, e.g. [38, 39], to explore these questions.

One of the methods we identified as appealing due to its intimate connection with the optimization procedure we use is that of [40]. There, a sensitivity matrix with respect to parameters (and possibly initial conditions) is leveraged to construct the Hessian of the variation of the objective function, in our case the Euclidean norm, with respect to model parameters. Inversion of the Hessian leads to the confidence intervals associated with each of the parameters (see, e.g., Eq. (6) therein). When we carried out this program, we found that it was not possible to invert the Hessian in a meaningful way because it was singular: the Hessian possesses two eigendirections associated with zero eigenvalues. By this program above, we mean in more detail the following: assume that the variables, the populations in the ODE model, are denoted by **x**, the parameters by ***θ***, and the dynamical equations Eqs. (1-9) by *x*_*i*_ = *f*_*i*_(**x, *θ***) for *i* = 1, 9. Without loss of generality we consider the optimizations with *α*_1_ = 0, *α*_2_ = 1, where the norm Eq. (12) depends only on *D*_num_(*t*) = *D*(*t*) with *D*(*t*) ≡ *x*_9_ in the dynamical equations. The Hessian, the quantity of interest, evaluates to

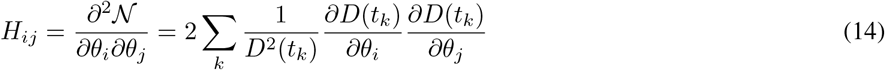

where the second equation arises from the explicit differentiation of the Euclidean norm coupled to the Gauss-Newton approximation, which neglects the second order derivative terms in the sum. This approximation is reasonable for second order derivatives smaller than the first order or for models that fit the observations well. Inspection of the Hessian suggests the connection between its eigenvalues and the confidence intervals. For small, tending to zero, eigenvalues large variations of the associated parameters do not alter significantly the (optimized) norm rendering them non-identifiable. The Hessian may be expressed in terms of the sensitivity matrix

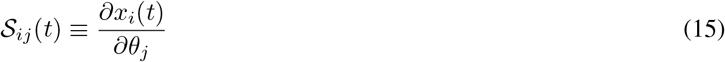

(where *i* enumerates variables, while *j* enumerates parameters) to become

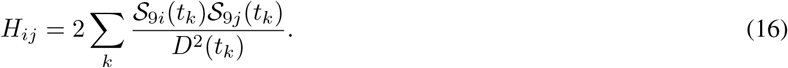

In the latter expression both *i* and *j* enumerate parameters. Hence, the determination of the Hessian reduces to the calculation of the sensitivity matrix. From the expression for the dynamical equations of motion, it is possible to formulate a differential equation which reads (component-wise):

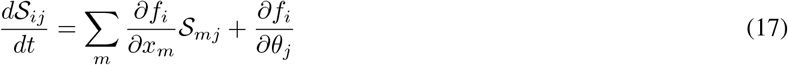

Again here *i* and *m* enumerate the variables, while *j* enumerates the parameters. This equation is integrated *together* with the equations of motion and at the final time and for the parameters of interest, it is used together with Eq. (16) to evaluate the Hessian matrix. For further details on the general approach, the reader is referred to [40]. It is this matrix that has been evaluated and its eigendirections have been identified. Notice that here the sensitivity with respect to parameters has been assessed; it is also, in principle, possible to assess using this set of tools, the sensitivity with respect to initial condition variations, although this has not been our focus herein. This aspect (of the 2 singular eigendirections of the Hessian) is connected with the identifiability of the model. Indeed, inspection of the model suggests that *ω, χ* and *ψ* are not independent parameters, but rather it is suitable to lump *ωψ* and (1 − *ω*) *χ* into just two lumped parameters. Alternatively, one can fix *ω* and optimize *ψ* and *χ*. This reduces the dimensionality of the kernel of the Hessian by one; however, it was not possible to identify the additional singular direction (e.g., of the corresponding Hessian eigenvector) by inspection.

Our experience with the model, as well as the analysis of [41] that considered reductions of the parameters of a similar model into irreducible combinations, suggested that the combinations *β*_*SA*_*ϕ* and *β*_*SI*_ (1 − *ϕ*) might be relevant to identifiability issues. These parameter combinations arise naturally when the model is rendered non-dimensional, in addition to appearing explicitly in the expression for the basic reproduction number Eq. (13). They determine the number of individuals exposed to the virus within the model, and hence there is a trade off between their respective contribution to the exposed class of individuals which, in turn, leads to the potential occurrence of fatalities. In that vein, we can expect an inverse relation between their relative magnitude, i.e., when the contribution of asymptomatics dominates the emergence of exposed, *β*_*SA*_*ϕ* will be larger than *β*_*SI*_ (1 − *ϕ*), while the opposite scenario should be possible (as well as scenarios “in between”). This would effectively amount to a “singular” direction of relative magnitude in the emergence of viral exposures. To test this assumption, we performed the optimizations reported in this section: the results are shown in Fig. 6.

**FIG. 6.**
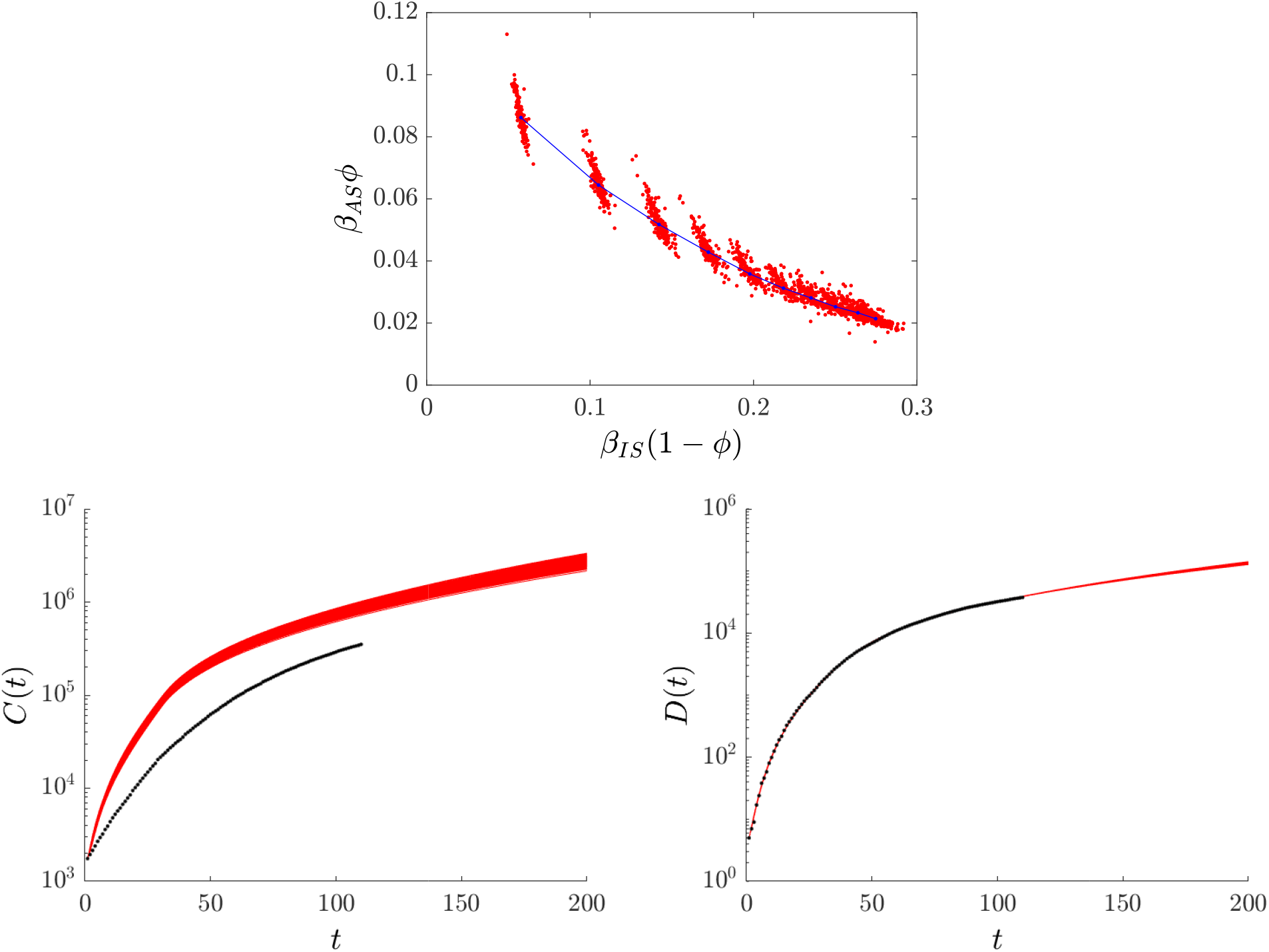
The top panel illustrates the inverse relationship between *β*_*SA*_*ϕ* and *β*_*SI*_ (1 − *ϕ*) for a wide range of optimizations performed under different constraints on the ratio *β*_*SA*_*/β*_*SI*_. Model parameters and initial conditions were varied. Red dots correspond to the optimal parameter values, blue dots are the median of the optimized values for each *β*_*SA*_*/β*_*SI*_ ; the line is a guide for the eye. The bottom panel show the curves of fits of cases (left) and fatalities (right) and associated predicted forward projections for all the parametric blobs of the top panel. As before, black denotes observation, while red represents the model predictions.

For the optimizations discussed in this section, section II C we varied both model parameters and initial conditions (in fact, the ratios *E*(0), *A*(0), *P* (0) to *I*(0)) as in Section II B. Similarly, the range of parameter variations was as specified in Table I. Distinctly from the optimizations of the previous section, we imposed an additional constraint on the relative magnitude of the two transmission rates, i.e., on *β*_*SA*_*/β*_*SI*_. We chose the relative magnitude to take ten equidistant values from 0.2 to 2.0. Each blob in top panel of Fig. 6 corresponds to a different ratio, the easiest to identify blobs (on the left) correspond to smaller *β*_*SA*_*/β*_*SI*_ ratios. Each blob presents the results of 200 optimization runs. The evolution of the entire set of 2000 runs as regards the predictions for the number of fatalities *D*(*t*) and cases *C*(*t*) is presented in the bottom panels of Fig. 6.

It is important to note that in *all* cases of the different blobs, the residual of the optimization was found to be comparable. This, practically, means that if we were considering any isolated form of each of these parameters, the corresponding confidence intervals would be vast and not particularly informative, due to the singular eigendirection of the Hessian. Nevertheless, this wide range of variation provides an equally adequate predictor of the time series for the fatalities. Hence, a far more meaningful question is to assess the potential variation of the projected number of fatalities in the future as a result of this “flexibility” (or invariance or implied lack of structural identifiability) of the model.

Inspection of Fig. 6 provides support to our claim that an inverse relation between the two combinations exists and that, in addition, a singular direction effectively exists within the problem. As a consequence, the plot of fatalities on the right panel illustrates convincingly, in our view, that the above uncertainty (in model parameters and initial conditions) does not have a major bearing in the variation of the model-predicted number of fatalities. This shows that while the specific parameters of our optimization may only be indicative of the order of magnitude of the associated quantities, within the wide range of freedom enabled by these singular directions, nevertheless, this (as well as the lack of structural identifiability of our model) does not significantly impact the reliability of our forward predictions. Similar issues naturally arise in the two-age model, and the principal ideas considered herein apply in such extensions of the model as well.

## III. TWO-POPULATION MODEL

### A. Equations

We now turn to the two-population variant of the model. Recall that due to the different characteristics of the two populations as considered herein, below and above 65 years, we expect that this model will be more adequate in capturing both deaths and cumulative infections of the full population. This is because on the one hand, the younger population in our considerations is more active (belonging typically to the workforce), hence has a different number of contacts. On the other hand, the older population has its own vulnerabilities to the virus SARS-CoV-2. As explained in the Introduction, the prevalence of various risk factors within the Mexican population [24] renders this partition even more relevant towards capturing the detailed data trends.

Firstly, we discuss the mathematical structure of the model. Here, we assume that each of the populations has its own set of parameters. Superscript *y* will denote parameters associated with the younger population, while superscript *o* will be connected to the older population. Naturally, the number of variables (*S, E, A, P, I, H, R, D*) now doubles, with each part having a younger and an older component. Note that some quantities that are associated with the virus, such as *σ*_1_ and *σ*_2_, are taken to be independent of age. Lastly, we explore a *mildly anisotropic* variant of the model whereby *β*^*o*^ ≡ *β*^*oy*^ = *β*^*yo*^ = *β*^*oo*^ and *β*^*y*^ ≡ *β*^*yy*^. That is to say, we assume that the older population has a different interaction (transmissions rate) with itself and with the younger individuals than the younger members of the population with themselves [19]. This is a reasonable assumption since the more sensitive older members of the population are advised to reduce their interactions, but they may be more infectious (higher viral load). While, in principle, we could have used a fully anisotropic variant of the model, we prefer to reduce the overall number of model parameters.

The differential equations for the two-age model are

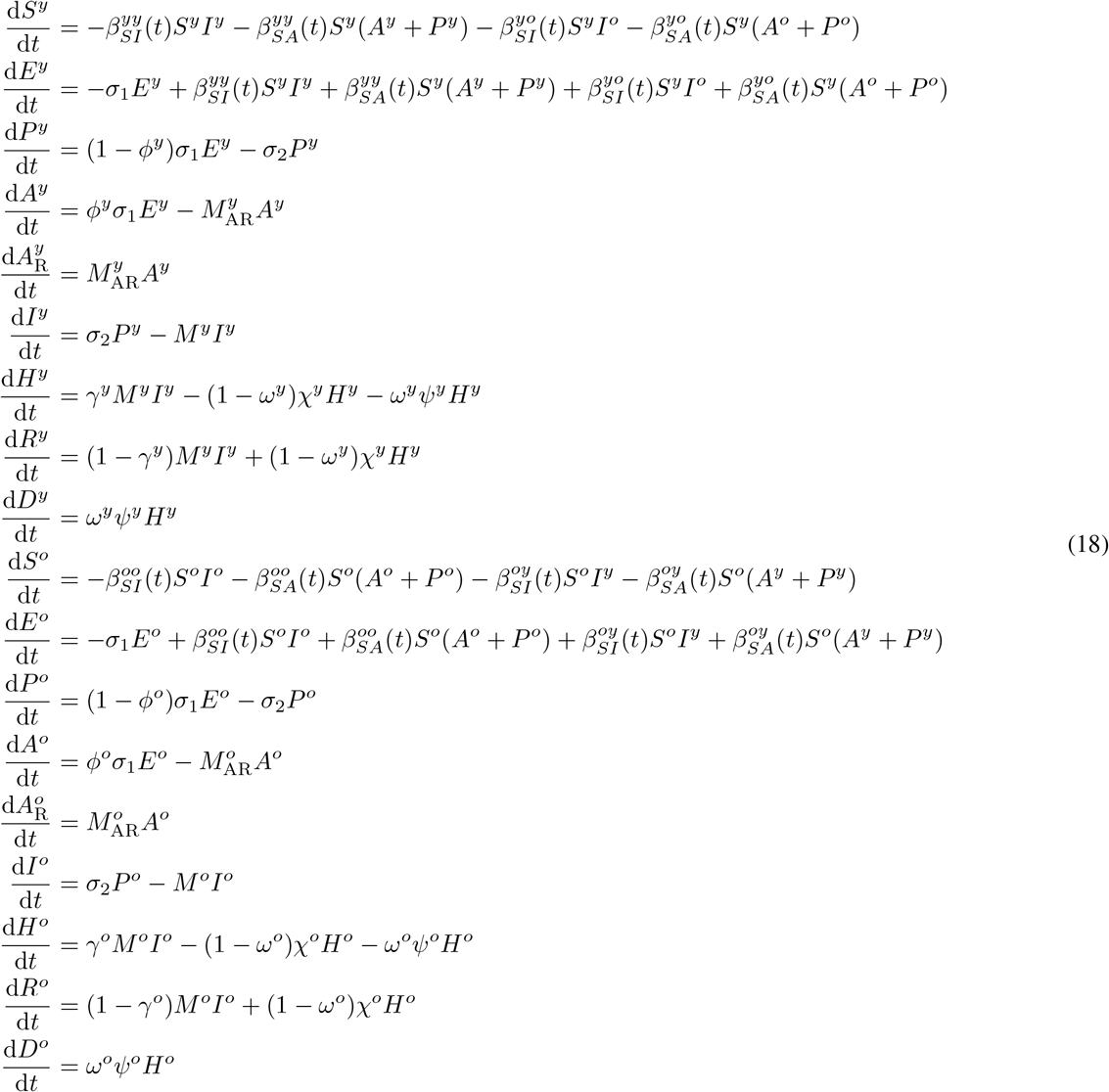

In a natural extension of what we assumed in the single-population model, we consider that as a result of the actual (imposed) partial mobility restrictions (social distancing) the temporal variation of the transmission rates *β*’s is:

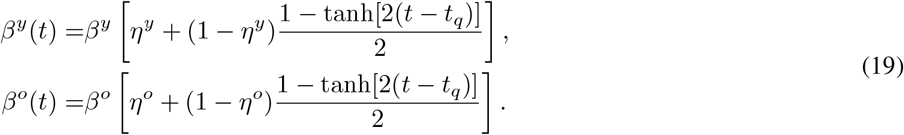

This implies that each of the *β*’s (associated with younger or older populations) was reduced by a respective factor of *η* at *t* = *t*_*q*_. Recall that part of our motivation is to assess the impact of additional intervention measures in the form of stricter mobility restrictions (lockdown) and the associated reduction of contacts, as well as the required use of personal protective equipment, such as face masks. In that light, for each one of the populations, we assume again that infected members maintain their *β*_*SI*_’s (in their quarantine modified by *η*_*SA*_), while the circulating asymptomatic (and presymptomatic) individuals would be affected by these measures through a further reduction, beyond the reduction due to the *η*’s, of their *β*_*SA*_’s by factors *ζ*^*y*^ and *ζ*^*o*^. The associated temporal dependence of the transmission rates then reads:

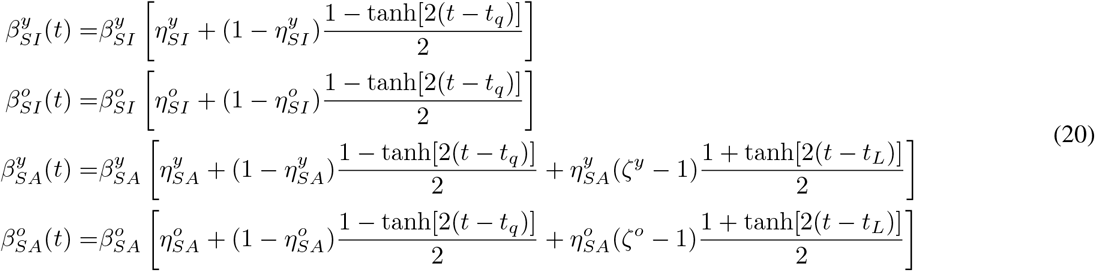

Figure 7 shows the time dependence of the transmission rates. for the scenario in which *only* the old population is required to adhere to the additional restrictive measures (suggested to be imposed as of August 10), i.e. *ζ*^*y*^ = 1 and *ζ*^*o*^ ≡ *ζ*. This is intended to explore the mitigation of contacts within the older population alone. In other cases, such an approach has been argued to be a reasonably safe approach towards attempting to return to economic and social stability [42]. Later, we will explore the possibility of lockdown measures imposed on both older and younger populations. The latter will be more relevant to compare with the single-population results of the previous section.

**FIG. 7.**
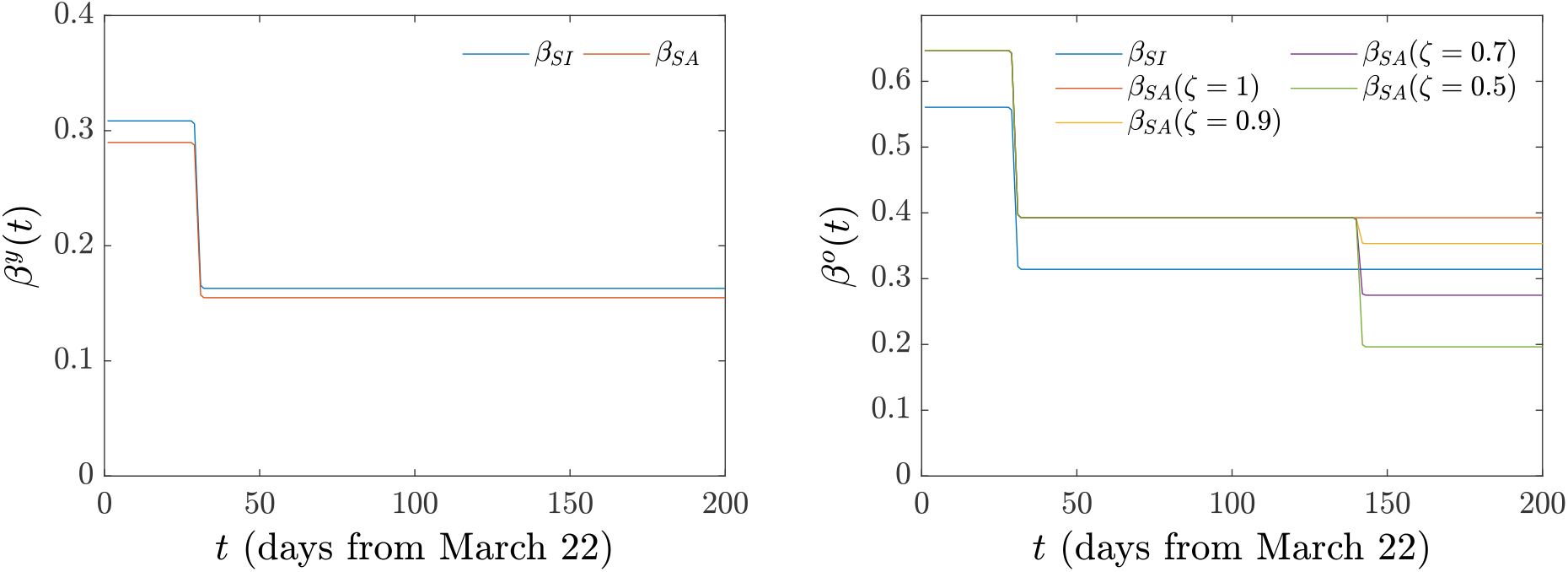
Two-population model. Time dependence of transmission rates *β*^*y*^, *β*^*o*^ under the existing policy (*ζ* = 1) and under the effect of additional (suggested) lockdown measures if applied solely to the older (than 65) population, i.e. *ζ*^*o*^ = 1 and *ζ*^*y*^ ≡ *ζ*.

As regards the aspect of identifiability of the age-structured model, we note the following. Importantly, and in order to reduce the number of relevant parameters, as indicated above, we have assumed that there are only two sets of *β*’s (rather than the potential four). Moreover, the characteristics pertaining to the virus itself, namely 1*/σ*_1_ and 1*/σ*_2_, associated, respectively, with the latent and preclinical periods, are assumed to be independent of age group. As a result of these selections, in this case of the two age groups, the *unknown parameters per age group* are fewer than in the case of a single age group. On the other hand, the number of data points involved in the corresponding optimization is indeed doubled, since we will be incorporating in the procedure the data for both the younger and the older population. It is relevant to also note that once again, we use the age-partitioned data in order to identify reasonable estimates of the fatality fraction *ω* to fix in the case of each of the age groups.

## B. Results

We follow an optimization procedure similar to the single-population case, with the initial values for the parameters within the ranges reported in Table III. As before, 2,000 optimizations were performed, parameters were uniformly sampled, and the ratio of initial conditions was included in the optimization. We have confirmed that in the case of 3,000 and 4,000 optimizations, the results are essentially identical to what is reported below. The optimal parameters are found by minimizing the norm

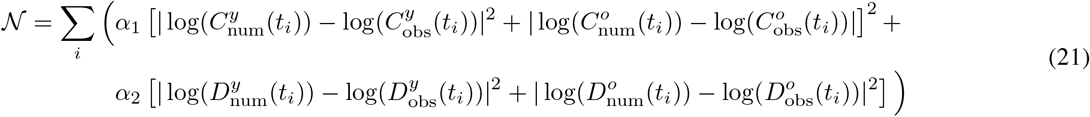

**TABLE III.** Two-populations model. Optimal model parameters (median and interquantile range). Euclidean-norm minimization with *α*_1_ = 0, *α*_2_ = 1. Fixed fraction of hospitalized who died *ω*^*y*^ = 0.0991, *ω*^*o*^ = 0.3248 (see text for justification). Variation range used in the optimization algorithm (initial parameter and initial-condition guesses were uniformly sampled within the ranges shown).

| Parameter | Symbol | Median (interquartile range) | Initial value |
| --- | --- | --- | --- |
| Population | $N$ | 127,575,528 | |
| Initial populations (young) | $(S_0^y; I_0^y; H_0^y; R_0^y; A_{R_0}^y; D_0^y)$ | (122,472,507 ($S_0^y/N = 0.96$); 1,370; 132; 2; 0; 3) | |
| Initial populations (old) | $(S_0^o; I_0^o; H_0^o; R_0^o; A_{R_0}^o; D_0^o)$ | (5,103,021 ($S_0^o/N = 0.04$); 207; 39; 2; 0; 2) | |
| Transmission rate ($y$) [per day] | $\beta_{SI}^y$<sup>a</sup> | 0.3085 (0.2862–0.3295) | $c \in U[0, 1]$ |
| Transmission rate ($y$) [per day] | $\beta_{SA}^y$<sup>a</sup> | 0.2897 (0.2725–0.3126) | $c \in U[0, 1]$ |
| Transmission rate ($o$) [per day] | $\beta_{SI}^o$<sup>a</sup> | 0.5605 (0.5419–0.5796) | $c \in U[0, 1]$ |
| Transmission rate ($o$) [per day] | $\beta_{SA}^o$<sup>a</sup> | 0.6471 (0.6297–0.6625) | $c \in U[0, 1]$ |
| Social distancing ($y$) effect | $\eta_{SI}^y$ | 0.5283 (0.5110–0.5458) | $c \in U[0, 1]$ |
| Social distancing ($y$) effect | $\eta_{SA}^y$ | 0.5345 (0.5121–0.5533) | $c \in U[0, 1]$ |
| Social distancing ($o$) effect | $\eta_{SI}^o$ | 0.5606 (0.5407–0.5790) | $c \in U[0, 1]$ |
| Social distancing ($o$) effect | $\eta_{SA}^o$ | 0.6068 (0.5876–0.6250) | $c \in U[0, 1]$ |
| Latent period [days] $\tau_l$ | $1/\sigma_1$ | 2.9699 (2.9673–2.9727) | $1/k, k \in U[2.75, 3.25]$ |
| Preclinical period [days] $\tau_p$ | $1/\sigma_2$ | 2.4695 (2.4678–2.4711) | $1/k, k \in U[2.25, 2.75]$ |
| A/P partitioning ($y$) | $\phi^y$ | 0.3710 (0.3405–0.3996) | $c \in U[0, 1]$ |
| A/P partitioning ($o$) | $\phi^o$ | 0.2581 (0.2442–0.2740) | $c \in U[0, 1]$ |
| Infectivity period (I, $y$) [days] $\tau_{inf}^{I,y}$ | $1/M^y$ | 4.1815 (4.1167–4.2442) | $1/k, k \in U[3, 14]$ |
| Infectivity period (I, $o$) [days] $\tau_{inf}^{I,o}$ | $1/M^o$ | 6.6060 (6.4495–6.7548) | $1/k, k \in U[3, 14]$ |
| Infectivity period (A, $y$) [days] $\tau_{inf}^{A,y}$ | $1/M_{AR}^y$ | 7.1888 (7.1049–7.2790) | $1/k, k \in U[5, 14]$ |
| Infectivity period (A, $o$) [days] $\tau_{inf}^{A,o}$ | $1/M_{AR}^o$ | 7.3453 (7.3183–7.3748) | $1/k, k \in U[5, 14]$ |
| Conversion fraction (I to H, R, $y$) | $\gamma^y$ | 0.2919 (0.2874–0.2965) | $c \in U[0.1, 0.4]$ |
| Conversion fraction (I to H, R, $o$) | $\gamma^o$ | 0.6438 (0.6370–0.6499) | $c \in U[0.45, 0.85]$ |
| Recovery period (H to R, $y$) [days] | $1/\chi^y$ | 7.8047 (7.6836–7.9446) | $1/k, k \in U[5, 15]$ |
| Recovery period (H to R, $o$) [days] | $1/\chi^o$ | 11.0976 (10.9733–11.2296) | $1/k, k \in U[7, 21]$ |
| H to D period ($y$) [days] | $1/\psi^y$ | 6.8216 (6.7721–6.8818) | $1/k, k \in U[5, 15]$ |
| H to D period ($o$) [days] | $1/\psi^o$ | 10.7787 (10.6725–10.8327) | $1/k, k \in U[5, 15]$ |
| Initial population fraction (E, $y$) | $E^y(0)/I^y(0)$ | 2.0459 (2.0077–2.1059) | $c \in U[1, 3]$ |
| Initial population fraction (E, $o$) | $E^o(0)/I^o(0)$ | 1.9938 (1.9443–2.0433) | $c \in U[1, 3]$ |
| Initial population fraction (A, $y$) | $A^y(0)/I^y(0)$ | 2.0481 (2.0111–2.1125) | $c \in U[1, 3]$ |
| Initial population fraction (A, $o$) | $A^o(0)/I^o(0)$ | 2.0085 (1.9651–2.0669) | $c \in U[1, 3]$ |
| Initial population fraction (P, $y$) | $P^y(0)/I^y(0)$ | 0.5487 (0.5316–0.5677) | $c \in U[0, 1]$ |
| Initial population fraction (P, $o$) | $P^o(0)/I^o(0)$ | 0.4481 (0.4284–0.4669) | $c \in U[0, 1]$ |

<sup>a</sup> The reported transmission rates $\beta$ have been multiplied by $N$ . They should be normalized by the initial population when used in ODE model.

with *α*_1_ = 0, and *α*_2_ = 1. Results are displayed in Fig. 8. Table III presents the optimal model parameters in terms of their median and the associated interquartile range, as well as the variation interval of the initial guesses of the parameters and the initial conditions (uniform sampling within each interval). We note that, as in the case of the single-population model, the fraction of hospitalized who died for the younger and older groups were fixed at *ω*^*y*^ = 0.0991 and *ω*^*o*^ = 0.3248, respectively. We also performed the optimization with *α*_1_ = *α*_2_, finding (results not shown here for brevity) that it bears similar advantages, such as capturing more adequately *C*(*t*), and similar pathologies, such as overestimation of deaths, most notably in the younger population, but also in the older population.

**FIG. 8.**
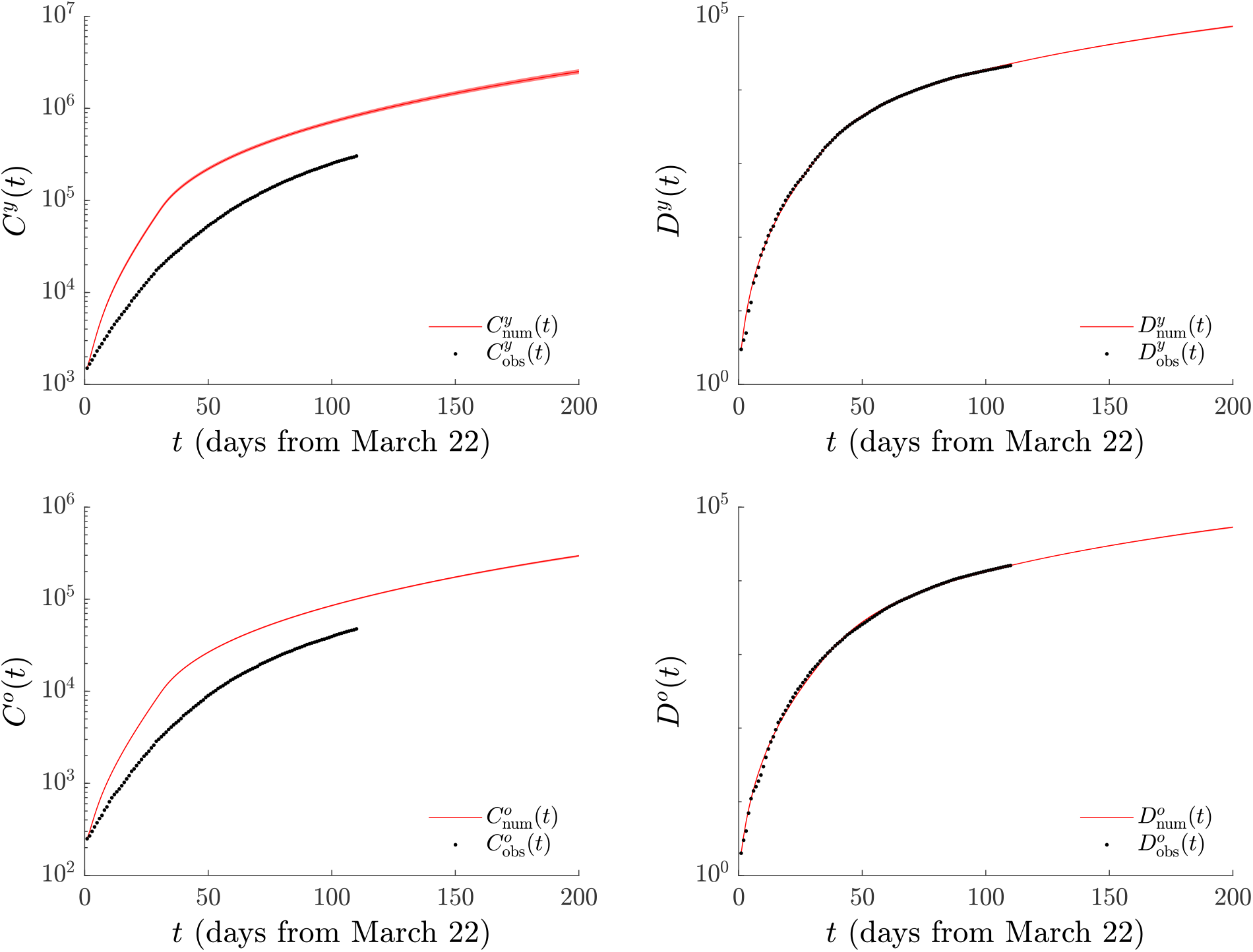
Two-population model. Number of cases (left) and of deaths (right) found by minimizing the norm (21) with *α*_1_ = 0, *α*_2_ = 1. The younger (than 65 years of age) population is shown in the top panels and the older (than 65) in the bottom panels. While the corresponding fatalities are somewhat comparable, recall that there is a far more significant susceptible population in the former category.

Comparison of the optimal (median) model parameters reported in Table III shows that some differences in the parameters for two populations (older and younger) persist. The transmission rates are considerably higher both for asymptomatic (presymptomatic) and infected individuals in the older population than in the younger population. We also note that within the symptomatic component of the respective populations, the younger one has a far smaller *γ* signifying a far smaller fraction of individuals that are symptomatically infected and need hospitalization. Once again, we highlight that one should not really interpret *ω* and *ψ* separately, but only the product thereof (which is still significantly larger for the older population) and similarly not consider (1 − *ω*) and *χ* separately, but once again their product which suggests a significantly larger recovery rate for the younger population.

We now turn to the comparison of the implications of maintaining *ζ* = 1 (i.e., current policy) vs. the projection upon a reduction of *ζ* by a factor of 0.9 to 0.5. The relevant results can be found in Table IV and Fig. 9. It is evident that a reduction of *ζ* through more severe mobility restrictions (lockdown) or behavioral interventions (wearing personal protective equipment) reduces according to the model the number of infections by anywhere between nearly 26 to about 112 thousand in the two cases. The respective projections for loss of human life range from about 680 (in the former case) to over 3,100 people (in the latter case) in this time-interval of a month. Clearly, the predictions of the model, both for the interval of August 10 to September 10 that is specifically monitored, but also over the longer time scale presented in Fig. 9 seem to warrant the consideration of such measures to the degree possible.

**TABLE IV.** Two-population model. Predictions for *C* and *D* at September 10 for parameters found by fitting the norm (21) and with additional measures applied to the older (than 65) population, i.e. *ζ*^*o*^ = 1 and *ζ*^*y*^ ≡ *ζ*.

| | $\zeta = 1$ | $\zeta = 0.9$ | $\zeta = 0.7$ | $\zeta = 0.5$ |
| --- | --- | --- | --- | --- |
| $C^y$ (total) | 1,883,242 (1,784,282–1,992,226) | 1,863,709 (1,763,823–1,970,731) | 1,827,439 (1,729,200–1,931,813) | 1,796,600 (1,698,648–1,898,648) |
| $C^o$ (total) | 223,824 (218,509–227,798) | 218,728 (213,482–222,682) | 209,257 (204,291–213,066) | 200,816 (195,949–204,414) |
| $C^y + C^o$ (total) | 2,107,241 (2,005,525–2,216,546) | 2,081,926 (1,981,237–2,191,014) | 2,035,929 (1,936,897–2,142,429) | 1,996,385 (1,899,808–2,101,006) |
| $C^y$ (from Aug. 10) | 581,179 (542,325–619,466) | 561,162 (523,067–597,982) | 524,178 (487,942–560,019) | 492,992 (458,648–527,456) |
| $C^o$ (from Aug. 10) | 68,227 (65,865–70,046) | 63,101 (60,889–64,934) | 53,675 (51,612–55,287) | 45,185 (43,247–46,669) |
| $C^y + C^o$ (from Aug. 10) | 649,616 (608,525–688,941) | 623,784 (584,123–663,089) | 577,908 (540,169–615,545) | 537,812 (502,401–574,215) |
| $D^y$ (total) | 54,139 (52,909–55,101) | 53,853 (52,629–54,811) | 53,331 (52,123–54,275) | 52,866 (51,673–53,809) |
| $D^o$ (total) | 39,423 (38,658–39,957) | 39,027 (38,282–39,560) | 38,288 (37,555–38,797) | 37,590 (36,881–38,091) |
| $D^y + D^o$ (total) | 93,595 (91,549–95,052) | 92,924 (90,901–94,347) | 91,628 (89,687–93,081) | 90,474 (88,556–91,901) |
| $D^y$ (from Aug. 10) | 17,566 (16,888–18,080) | 17,277 (16,615–17,791) | 16,751 (16,109–17,260) | 16,301 (15,672–16,800) |
| $D^o$ (from Aug. 10) | 12,991 (12,538–13,304) | 12,600 (12,157–12,910) | 11,855 (11,436–12,154) | 11,160 (10,760–11,456) |
| $D^y + D^o$ (from Aug. 10) | 30,559 (29,412–31,389) | 29,881 (28,762–30,700) | 28,617 (27,557–29,430) | 27,459 (26,451–28,262) |

**FIG. 9.**
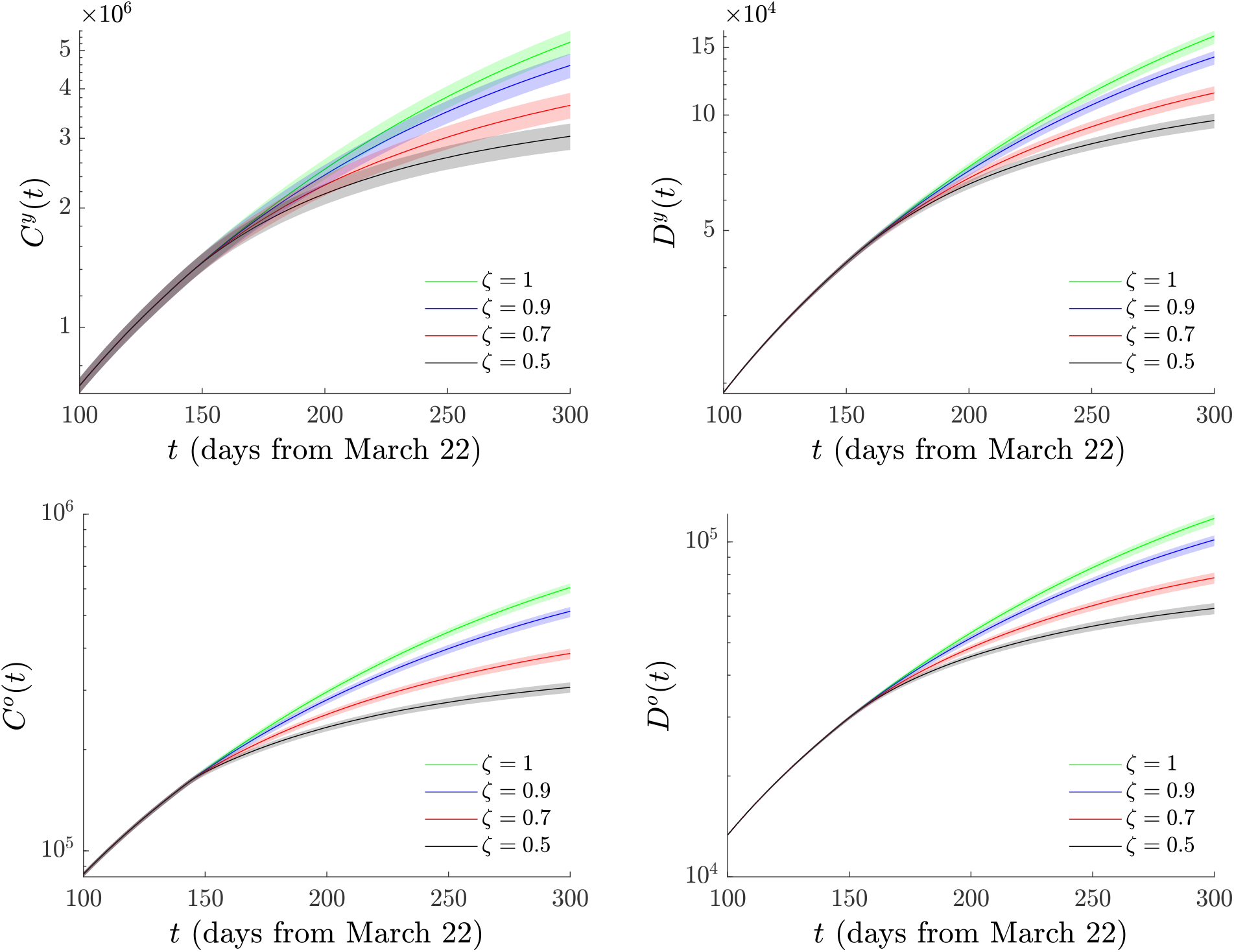
Same as Fig. 5, but for the two-population model when additional measures are applied to the older (than 65) population, i.e. *ζ*^*y*^ = 1 and *ζ*^*o*^ ≡ *ζ*. Notice the significant deviation of both the number of infections (left) and of the number of deaths (right), both for the younger (than 65) and the older (than 65) populations between current policy (*ζ* = 1) and the suggested additional intervention measures (by a factor of 0.5 to 0.9).

Let us now touch upon the case when intervention measures are applied to both populations. The corresponding variation of the *β*’s involving an equal restrictive factor *ζ* for both young and old populations is shown in Fig. 10. In turn, Table V shows the effect of these restrictive measures applied equally to both populations on both deaths and cumulative infections (compared to Table II where intervention measures were applied to the single-age, full population model). The predicted development of the pandemic refers to the period from August 10 to September 10, i.e., for one month. Figure 11 presents a longer time-scale perspective of these mitigation effects, extending well past September 10.

**FIG. 10.**
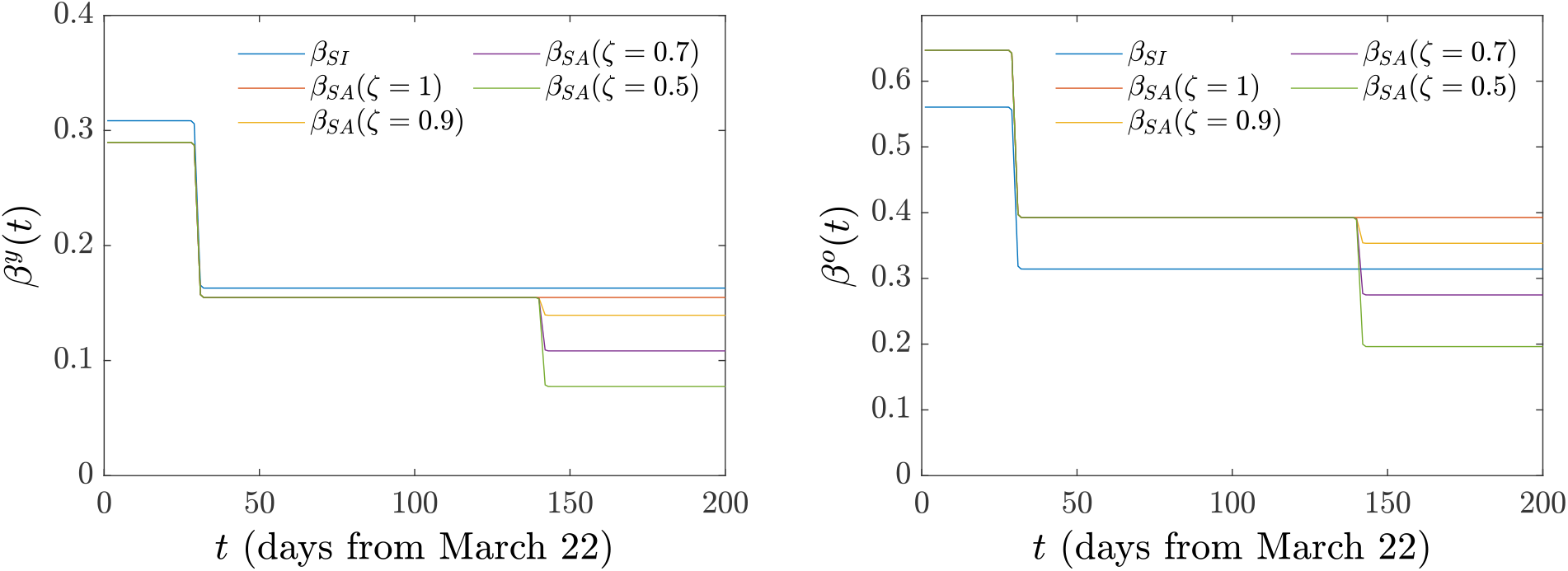
Two-population model. Time dependence of *β*^*y*^ and *β*^*o*^ under existing policy (*ζ* = 1) and under the effect of additional (suggested) lockdown measures when applied to both populations (i.e. *ζ*^*o*^ = *ζ*^*y*^ ≡ *ζ*).

**TABLE V.** Two-population model. Predictions for *C* and *D* at September 10 for parameters found by fitting the norm (21) when the additional measures are applied to both populations (i.e., *ζ*^*o*^ = *ζ*^*y*^ ≡ *ζ*).

| | $\zeta = 1$ | $\zeta = 0.9$ | $\zeta = 0.7$ | $\zeta = 0.5$ |
| --- | --- | --- | --- | --- |
| $C^y$ (total) | 1,883,242 (1,784,282–1,992,226) | 1,832,182 (1,734,273–1,936,665) | 1,743,348 (1,648,639–1,843,535) | 1,670,650 (1,578,768–1,765,596) |
| $C^o$ (total) | 223,824 (218,509–227,798) | 216,781 (211,645–220,702) | 204,799 (199,864–208,487) | 195,055 (190,424–198,644) |
| $C^y + C^o$ (total) | 2,107,241 (2,005,525–2,216,546) | 2,049,131 (1,948,776–2,154,878) | 1,948,409 (1,851,052–2,048,446) | 1,864,747 (1,772,443–1,961,328) |
| $C^y$ (from Aug. 10) | 581,179 (542,325–619,466) | 529,226 (493,315–566,658) | 439,465 (407,489–471,540) | 366,302 (338,115–395,001) |
| $C^o$ (from Aug. 10) | 68,227 (65,865–70,046) | 61,211 (58,966–62,932) | 49,161 (47,234–50,684) | 39,416 (37,737–40,897) |
| $C^y + C^o$ (from Aug. 10) | 649,616 (608,525–688,941) | 590,436 (552,929–629,456) | 488,501 (455,084–521,819) | 405,910 (375,993–434,966) |
| $D^y$ (total) | 54,139 (52,909–55,101) | 53,373 (52,183–54,307) | 52,006 (50,858–52,889) | 50,826 (49,709–51,697) |
| $D^o$ (total) | 39,423 (38,658–39,957) | 38,926 (38,187–39,453) | 38,035 (37,319–38,540) | 37,263 (36,560–37,752) |
| $D^y + D^o$ (total) | 93,595 (91,549–95,052) | 92,331 (90,326–93,754) | 90,054 (88,153–91,443) | 88,074 (86,279–89,434) |
| $D^y$ (from Aug. 10) | 17,566 (16,888–18,080) | 16,793 (16,150–17,288) | 15,436 (14,831–15,894) | 14,245 (13,688–14,674) |
| $D^o$ (from Aug. 10) | 12,991 (12,538–13,304) | 12,492 (12,054–12,800) | 11,603 (11,186–11,897) | 10,820 (10,430–11,114) |
| $D^y + D^o$ (from Aug. 10) | 30,559 (29,412–31,389) | 29,301 (28,204–30,109) | 27,035 (26,025–27,795) | 25,063 (24,125–25,797) |

**FIG. 11.**
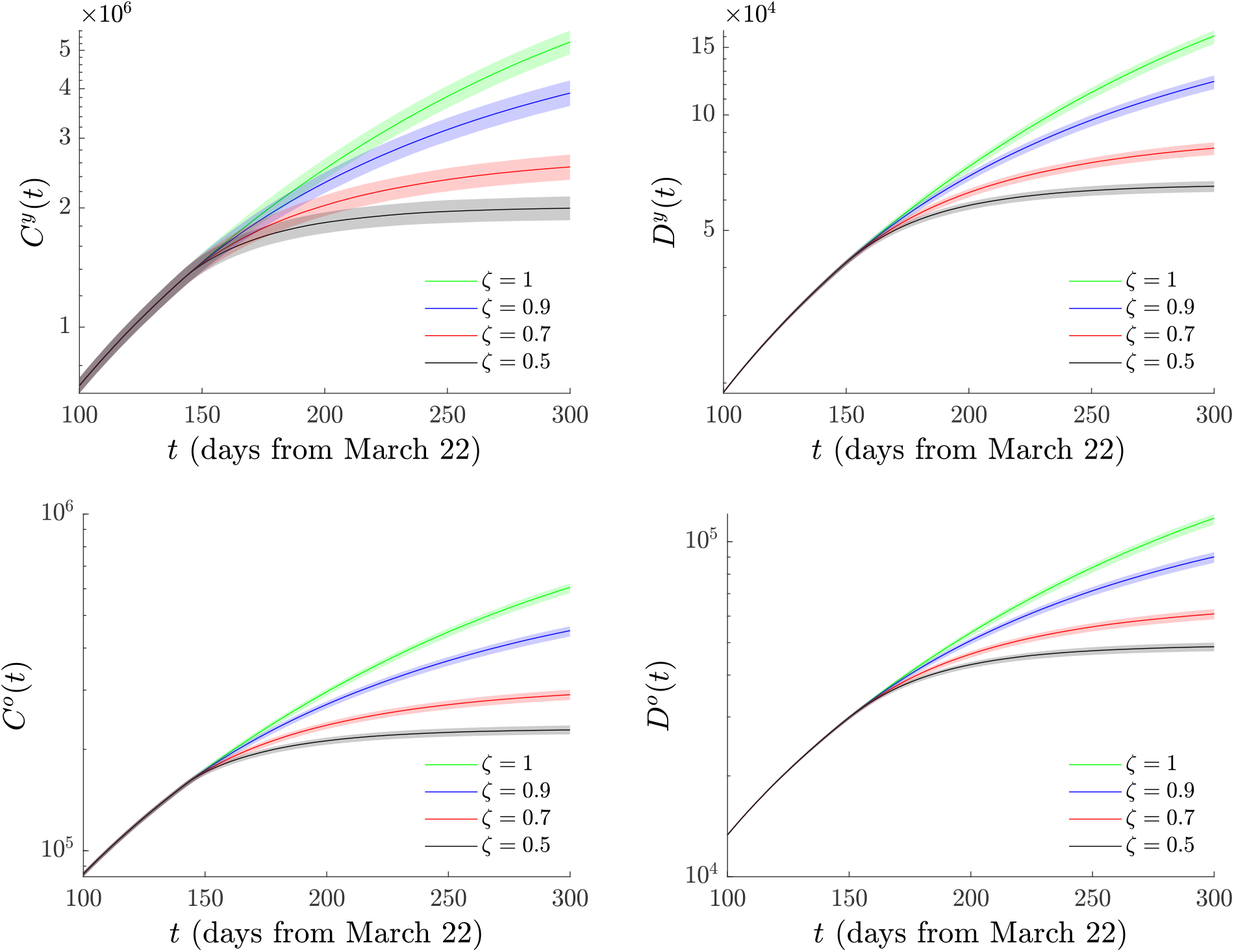
Same as Fig. 9 but now with the restrictive measures applied from August 10 onward to both populations (i.e. *ζ*^*o*^ = *ζ*^*y*^ ≡ *ζ*, transmission rates as depicted in Fig. 10).

A complementary view of the potential effect of measures can be observed in Fig. 12. Here the number of potentially fewer deaths as predicted by the model is presented both in the case of the single-age model, as well as in that of the two-age model, both for restriction of just the older population (red curve, left panel) and of the older and younger populations (blue curve, left panel). It can be seen that the age-structured two-population model predicts fewer avoided deaths with respect to the less structured single-age mode. More concretely, from Table V, we notice that within the month of interest (August 10 to September 10), the decrease in infections is by around 58,000 for *ζ* = 0.9 or by close to 242,000 for *ζ* = 0.5. The decrease in number of deaths over the same time period is by about 1,260 for *ζ* = 0.9, while for *ζ* = 0.5 the decrease in deaths is by 5,496. We hope that these predictions of the model and the comparisons between restrictive measures for a single (older) population group vs. ones for the entire population, may be informative for public health considerations.

**FIG. 12.**
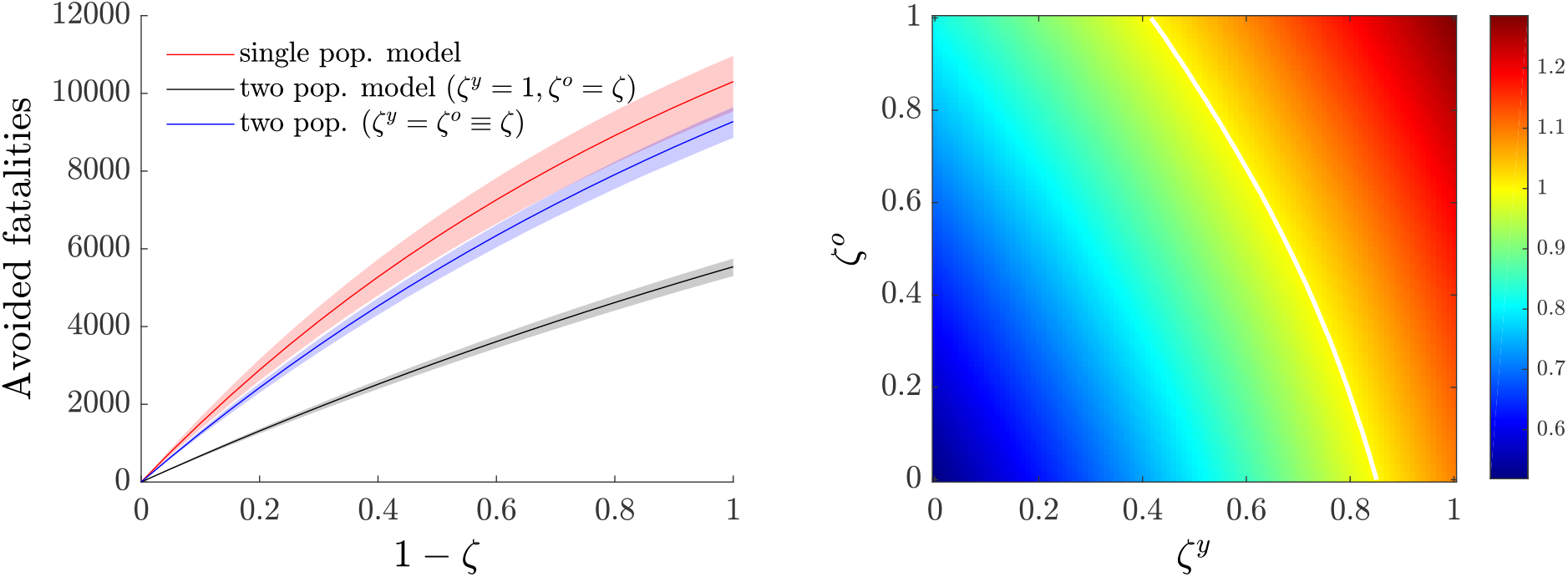
Number of fatalities that could have been avoided in the period Aug. 10 – Sept. 10 if the different measures discussed herein had been applied (left panel). Effective reproduction number (median) *R*_*e*_ as a function of *ζ*^*y*^ and *ζ*^*o*^ for the two-population model (right panel). The white solid curve corresponds to *R*_*e*_ = 1: hence, combinations of restrictive measures in the plane (*ζ*^*y*^, *ζ*^*o*^) to the left of the white curve are predicted to lead to subsiding of the pandemic, while above it they would lead to its growth.

We also calculated *R*_0_ for the two-population version of our model via

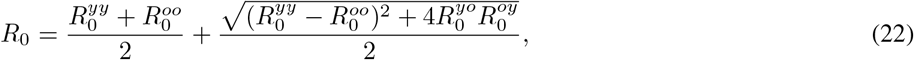

where the explicit expressions of 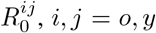, *i, j* = *o, y* are the given in Appendix A 2. We note, however, that 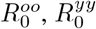, are identical to (13) in the case when the old and young groups, respectively, are isolated. 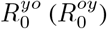 correspond to the case when the young (old) age group comes into contact with only old (young) infectious hosts. This form of community *R*_0_ has been found in other contexts, such in HIV spread using a multi-group model [43]. The reproduction number *R*_0_ and the effective reproduction number *R*_*e*_ at the beginning of social distancing measures (*t* = *t*_*q*_) for the optimized model parameters are given at Table VI. Upon imposition of the relevant measures, e.g., for *ζ* = 0.7 or *ζ* = 0.5, we have confirmed that both in the case of constraints only for the older population or in that of restrictive measures for both the older and the younger population, it is true that *R*_*e*_ < 1, as shown in right panel of Fig. 12. Indeed, the figure illustrates not only the scenarios of *ζ*^*y*^ = 1 and of *ζ*^*y*^ = *ζ*^*o*^ considered above, but also that of arbitrary restrictive combinations in the (*ζ*^*y*^, *ζ*^*o*^) plane and how they can lead to subsiding the pandemic (for *R*_*e*_ < 1, i.e., below the white curve).

**TABLE VI.**
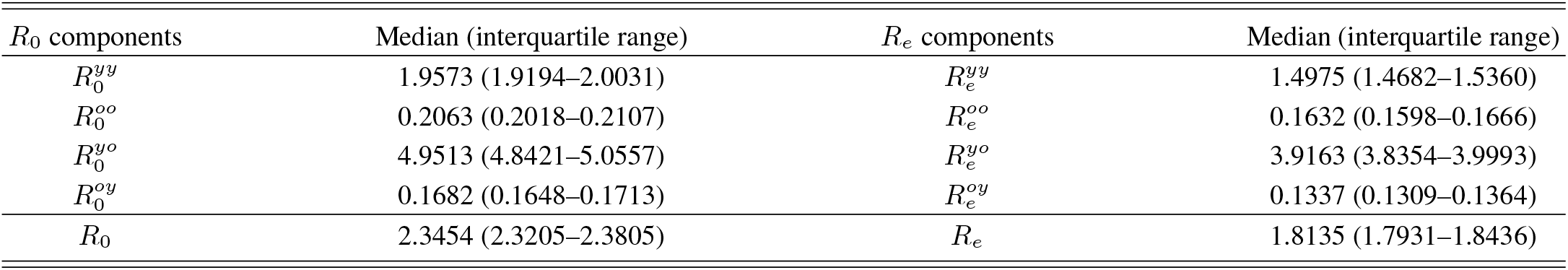
Reproduction number *R*_0_ and effective reproduction number *R*_*e*_ at the beginning of actual social distancing measures (*t* = *t*_*q*_) for the two-age model.

| $R_0$ components | Median (interquartile range) | $R_e$ components | Median (interquartile range) |
| --- | --- | --- | --- |
| $R_0^{yy}$ | 1.9573 (1.9194–2.0031) | $R_e^{yy}$ | 1.4975 (1.4682–1.5360) |
| $R_0^{oo}$ | 0.2063 (0.2018–0.2107) | $R_e^{oo}$ | 0.1632 (0.1598–0.1666) |
| $R_0^{yo}$ | 4.9513 (4.8421–5.0557) | $R_e^{yo}$ | 3.9163 (3.8354–3.9993) |
| $R_0^{oy}$ | 0.1682 (0.1648–0.1713) | $R_e^{oy}$ | 0.1337 (0.1309–0.1364) |
| $R_0$ | 2.3454 (2.3205–2.3805) | $R_e$ | 1.8135 (1.7931–1.8436) |

## IV. CONCLUSIONS AND FUTURE CHALLENGES

In the present work, we proposed a model for the examination and fitting of the progression of the pandemic of COVID-19, with an aim towards applications to data from different countries. The model was developed to include susceptible individuals, turning to exposed upon an infectious interaction. Subsequently, these are split to presymptomatic or asymptomatic after a latent period. The asymptomatics can only recover, while the presymptomatics are led to an onset of symptoms. Thereafter, this can lead to either recovery or the need for hospitalization, and the latter again can either lead to recovery or to fatality. The model naturally involves assumptions that are not always met. For example, in some cases people may die before getting a chance to be hospitalized. In other settings, where there is heavy testing involved, asymptomatics may be counted in the reported infections: while this is not an explicit assumption of the model, we did not count any fraction of asymptomatics in the cumulative infections when presenting the relevant comparison. Additionally, also, some of the hospitalized individuals may transmit the virus (e.g., to medical personnel) despite the much more substantial health and safety protocols applicable within hospitals. In any event, we consider these features to be the exception rather than the rule and hence have excluded them from our more mainstream considerations.

Within the realm of the model, we exposed the meaning of relevant parameters (e.g., transmission rates, latent and incubation times, fractions of asymptomatics vs. presymptomatics, of hospitalized vs. directly recovered, and of recovered vs. dying individuals at the hospital; also the time scales of the latter partitions were considered). Specifically, we attempted to assign an epidemiological meaning to our different parameters and to examine the associated results of the optimal fit of these parameters to the data from a specific time series to illustrate the “reasonable” nature of the findings. Notice that in addition to explaining the fitting process (to either deaths or deaths and cumulative infections), we took the approach of using a minimal number of assumptions to avoid over-constraining the system. We also illustrated how the model can be partitioned to different age groups, based on the data that may be available for the country or region of interest. Here, we opted to consider the simplest partition to two-age models. While tedious, it is structurally straightforward (and of some interest in its own right) to generalize considerations to many age group models.

As our prototypical illustration of choice, we used data from Mexico which became available through [27]. This is a case where a significant number of cases has arisen and the consideration of potential further lockdown measures is an important topic of ongoing debate. Indeed, our findings suggest that the present measures appear not to be sufficient to mitigate the catastrophic consequences of the pandemic, since the calculated current value of the effective basic reproduction number of the epidemic is *R*_*e*_ > 1 and hence the situation appears to need further mitigation measures and strategies to avoid significant loss of life. In that vein, we discussed in the realm of the model what consequences different additional intervention measures would have had at the level of deaths and of cumulative infections. We found, e.g., that a reduction of transmission rates by a factor of around 1*/*2 via social distancing or related measures (lockdown, requirement to wear personal protective equipment (e.g., face masks)) would lead to a nontrivial curbing of the rampant growth of both *C*(*t*) and *D*(*t*). The relevant reductions could be of the order of 100, 000 in terms of infections and of numerous thousands in terms of deaths in the interval of the 30 days subsequent to the data alone. While our results are only suggestive (and relevant within the context of the model), we hope that they may be of some value towards policy considerations in the near future.

Naturally, there are numerous directions that are worthwhile to consider towards extensions of the present work. A natural feature of many of the models (e.g., associated with the US [16], but also elsewhere) is the incorporation of uncertainty, beyond what we presented in this work. We are currently in the process of building into the formulation an uncertainty quantification framework on the basis of polynomial chaos considerations [44] and the use of suitable, non uniform (as in this work) distributions for quantities such as the transmission rates. Constructing a robust such framework would be of considerable value to models such as the one proposed herein. In addition, as indicated in numerous cases, data broken down by age groups exist, e.g., not only for Mexico, but also for other countries such as Portugal [45], etc. Clearly, a generalization of the model that considers the data by decade would offer a more complete and systematic picture of the impact of COVID-19 on different sub-populations and hence their potential risk. This would offer, in turn, a clearer picture of which age groups to attempt to protect and would be worthwhile (even if somewhat cumbersome). Lastly, as different countries are considering partial lockdowns or partial re-openings the formulation of a meta-population model with different hubs and a quantification of the transport coefficients between these [46] would be central towards going beyond the well-mixed assumption and factoring in a spatial structure and transportation features within the model. Such directions are currently under active investigation and will be reported in future work.

## Data Availability

All data are available upon request to authors

## ACKNOWLEDGMENTS

This material is based upon work supported by the US National Science Foundation under Grants No. DMS-1815764 (ZR), PHY-1602994, and DMS-1809074 (PGK). PGK also acknowledges support from the Leverhulme Trust via a Visiting Fellowship and thanks the Mathematical Institute of the University of Oxford for its hospitality during this work. PGK and ZR acknowledge support through the C3.ai Digital Transformation Institute.

*Disclaimer* The views expressed in this manuscript are purely those of the authors and may not, under any circumstances, be regarded as an official position of the European Commission.

## Appendix A Calculation of the reproduction number *R*_0_

### 1. Single-population model

We use the next-generation matrix approach to find *R*_0_ [37]. We define the relevant vectors:

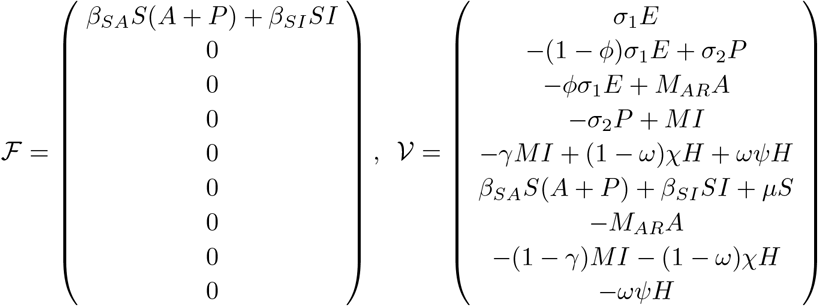

We then focus on the five infectious/infected compartments (*E, P, A, I, H*) and ignore the rest (*S, A*_*R*_, *R, D*). We find the Jacobians of *F, V* with respect to *E, P, A, I, H* in the order they appear. This yields two 5 × 5 matrices:

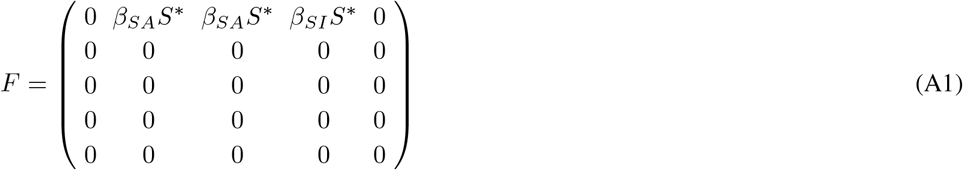

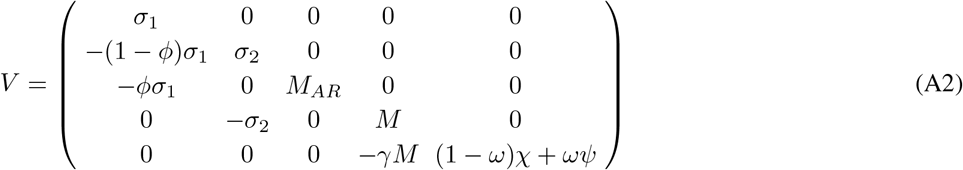

The basic reproduction number is the spectral radius of *FV* ^−1^ which in our case is

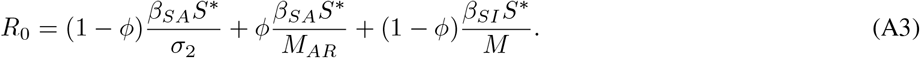

The first term is due to the presymptomatic hosts *P*, the second due to the asymptomatic hosts *A*, and the last one due to the symptomatic infectious hosts *I*. In each term, the numerator yields the rate of new infections *β*_*SA*_*S*^*^, *β*_*SI*_*S*^*^ and this is then multiplied with the average duration of the stay in that infectious stage *τ*_*p*_ = 1*/σ* _2_ (preclinical period), 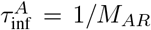 (asymptomatic infectious period), 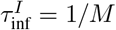 (symptomatic infectious period). Each term is multiplied by the fraction of hosts in that stage/state *ϕ* for asymptomatics and 1 − *ϕ* for symptomatics.

### 2. Two-population model

We use again the next-generation matrix approach as described in the previous Appendix A 1 to obtain the following two matrices:

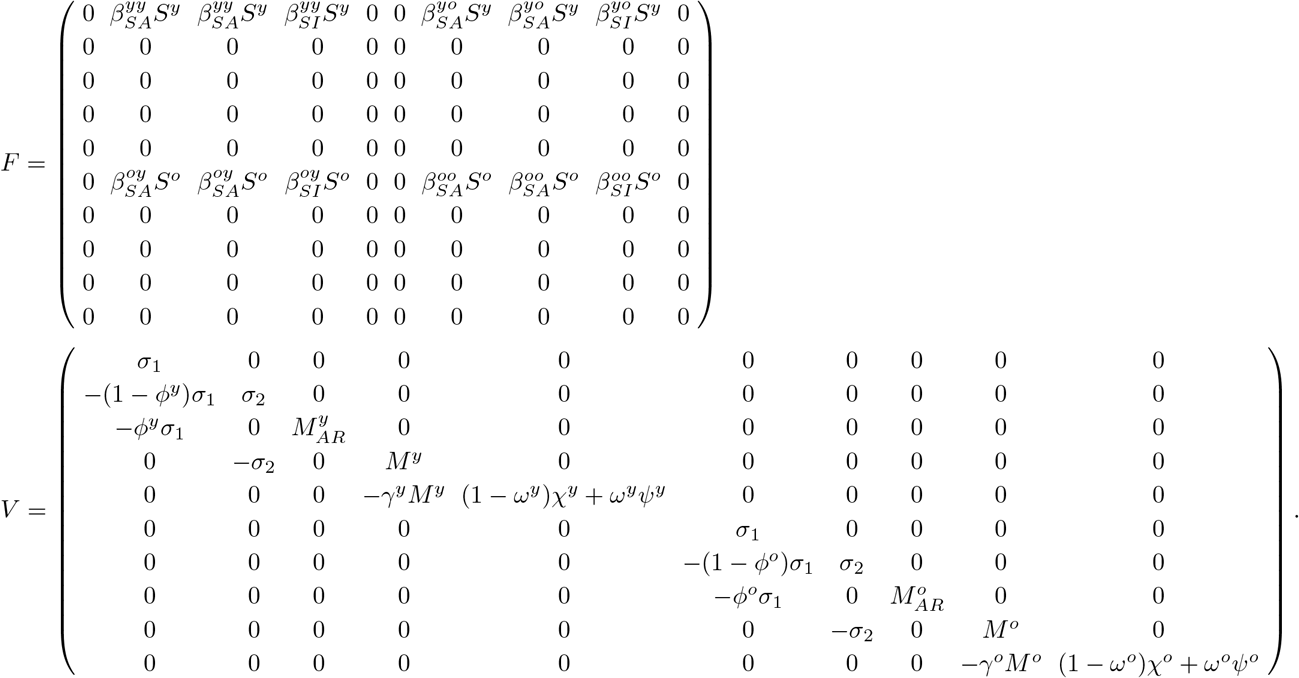

Since *V* is a block matrix it holds

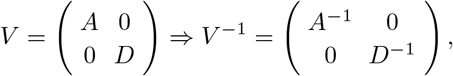

from which is readily follows that the spectral radius of *FV* ^−1^ is given by

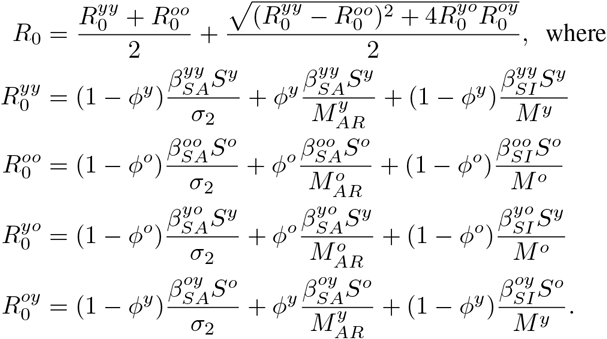

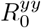 and 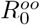 are the basic reproduction numbers in the young and old age groups, respectively, if they were completely isolated of each other. 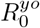 is the basic reproduction number if susceptible young hosts come into contact with only old infectious hosts, and 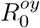 is the basic reproduction number if old hosts come into contact with only young infectious hosts.

## Notes

### Competing Interest Statement

The authors have declared no competing interest.

### Author Declarations

This field is not applicable in our case

## References

[1] https://www.who.int/emergencies/diseases/novel-coronavirus-2019

[2] W.O. Kermack and A.G. McKendrick, Proc. Roy. Soc. A 115, 700 (1927).

[3] H. Hethcote, SIAM Rev. 42, 599 (2000).

[4] N.T.J. Bailey, The mathematical theory of infectious diseases and its applications, Griffin (London, 1975).

[5] R.M. May and R.M. Anderson, Infectious diseases of humans: dynamics and control, doi:Oxford University Press (Oxford, 1991).

[6] F. Brauer and C. Castillo-Chávez, Mathematical Models in Population Biology and Epidemiology, doi:Springer-Verlag (New York, 2001).

[7] Z.-Q. Xia, J. Zhang, Y.-K. Xue, G.-Q. Sun, and Z. Jin, PLoS One 10, e0144778 (2015).

[8] X. Hao, S. Cheng, D. Wu, T. Wu, X. Lin, and C. Wang, Nature, https://doi.org/10.1038/s41586-020-2554-8 (2020).

[9] L. López and X. Rodó, Nat. Hum. Behav. 4, 746–755 (2020). https://doi.org/10.1038/s41562-020-0908-8

[10] M. M. Arons et al., Presymptomatic SARS-CoV-2 infections and transmission in a skilled nursing facility, N. Engl. J. Med., DOI: 10.1056/NEJMoa2008457.

[11] M. Peirlinck, K. Linka, F.S. Costabal, J. Bhattacharya, E. Bendavid, J.P.A. Ioannidis, and E. Kuhl, Comput. Methods Appl. Mech. Engrg. 372 113410 (2020).

[12] E. J. Emanuel et al., N. Eng.l J. Med. 382, 2049–2055 (2020).

[13] J. B. Dowd, L. Andriano, D.M. Brazel, V. Rotondia, P. Block, X. Ding, Y. Li, and M.C. Mills, Proc. Natl. Acad. Sci. U.S.A 117, 9696–9698 (2020).

[14] Z. Zheng et al., J. Infect. 81, e16–e25 (2020).

[15] M.R. Nepomuceno, E. Acosta, D. Alburez-Gutierrez, J.M. Aburto, A. Gagnon, and C.M Turra, Proc. Natl. Acad. Sci. U.S.A 117, 13881–13883 (2020).

[16] For instance in the models reported in: https://www.cdc.gov/coronavirus/2019-ncov/covid-data/forecasting-us.html for modeling the US, only one (Northeastern) appears to focus on age-structured modeling [see, e.g., https://covid19.gleamproject.org/ and references therein].

[17] J. E. Ludvigsson, Acta Paediatr.109 1088–1095 (2020).

[18] K. Xu et al., Clin.Infect. Dis. 71, 799–806 (2020).

[19] J. Mossong et al., PLoS Med 5, 374 (2008).

[20] M. A. Acuña-Zegarra, M. Santana-Cibrian, and J. X. Velasco-Hernandez, Math. Biosci. 325, https://doi.org/10.1016/j.mbs.2020.108370 (2020).

[21] O. Torrealba-Rodriguez R. A. Conde-Guttiérrez, A. L. Hernández-Javier, Chaos Solitons Fractals 138, 109946 (2020).

[22] M. Anzarut et al., https://arxiv.org/abs/2007.09117

[23] U. Avila-Ponce de León, A. G. C. Pérez, E. Avila-Vales, Big Data and Information Analytics, 5(1): 14–28 (2020). doi: 10.3934/bdia.2020002

[24] J. P. Gutierrez and S. M. Bertozzi, PLoS ONE 15(10): e0240394. https://doi.org/10.1371/journal.pone.0240394

[25] P.G. Kevrekidis, J. Cuevas-Maraver, Y. Drossinos, Z. Rapti, G.A. Kevrekidis, arXiv:2005.04527.

[26] See, e.g., https://elifesciences.org/articles/57309

[27] https://www.gob.mx/salud/documentos/datos-abiertos

[28] https://en.wikipedia.org/wiki/COVID-19_pandemic_in_Mexico

[29] N.I. Stilianakis and Y. Drossinos, J. R. Soc. Interface 7, 1355 (2010).

[30] Y. Drossinos and N.I. Stilianakis, Aerosol Sci. Technol. 54, 639 (2020).

[31] D.K. Milton, M. Patricia Fabian, B.J. Cowling, M.L. Grantham, J.J. McDevitt, PLoS Pathog. 9, e1003205 (2013).

[32] J.F. Robinson, I. Rios der Anda, F.J. Moore, J.P. Reid, R.P. Sear and C.P. Royall, arXiv:2008.04995v2

[33] A.A. King, M. Domenech de Cellés, F.M.G. Magpantay, P. Rohani, Proc. Roy. Soc. B 282, 20150347 (2015).

[34] https://ourworldindata.org/coronavirus/country/mexico?country=\~MEX

[35] See, e.g., the CDC report: https://www.cdc.gov/coronavirus/2019-ncov/cases-updates/commercial-lab-surveys.html and the associated discussion in: https://www.nytimes.com/2020/07/21/health/coronavirus-infections-us.html.

[36] M.C. Eisenberg, S.L. Robertson, J.H. Tien, J. Theor. Biol. 324, 84 (2013).

[37] O. Diekmann, J. A. P. Heesterbeek, and J. A. J. Metz, J. Math. Biol. 28, 365–382 (1990).

[38] K. Roosa, G. Chowell, Theor. Biol. Med. Model. 16: 1 (2019).

[39] G. Chowell, Infect. Dis. Model. 2, 379–398 (2017).

[40] T.G. Farmer Jr., Th. F. Edgar, N. Peppas, Diabetes Technol. Ther. 10, 128–141 (2008); T.G. Farmer Jr., Intravenous closed-loop glucose control in type I diabetes patients, Ph.D. thesis, University of Texas Austin (2007).

[41] A.S. Fokas, J. Cuevas-Maraver, P.G. Kevrekidis, Chaos Solitons Fractals 140, 110244 (2020).

[42] A.S. Fokas, J. Cuevas-Maraver, P.G. Kevrekidis, Sci. Rep. 11, 5839 (2021).

[43] J. M. Hyman and J. Li, SIAM J Appl. Math. 57 1082–1094 (1997).

[44] D. Xiu and G. Em. Karniadakis, SIAM J. Sci. Comput. 24, 619 (2002).

[45] https://github.com/dssg-pt/covid19pt-data

[46] R. Li et al., Science 368, 489 (2020).

